# Funding distributions, trends, gaps, and policy implications for spinal cord injury research: A systematic analysis of US federal funds

**DOI:** 10.1101/2025.06.01.25328764

**Authors:** Tucker Gillespie, Andrew Buxton, Bethany R. Kondiles, Miranda Leal-Garcia, Mia R. Pacheco, Ashley V. Tran, Katie Vo, Lucy Abu, James Barr, Tanya A. Barretto, Jason Biundo, Sam Duenwald, Abigail Evans, Timothy N. Friedman, Isabella Gadaleta, Saahas Ganesh, Bryson Gottschall, Peyton Green, Grant Lee, Lilian Liu, Raza N. Malik, Elizabeth J. Nava, Chiara Sorani, Vansh Tandon, Hannah Thomas, Kyndal Thomas, Chris Barr, Ian Burkhart, Dylan A. McCreedy, Peter Nowell, Heath Blackmon, Alexander G. Rabchevsky, Matthew Rodreick, Abel Torres-Espín, Jennifer N. Dulin

## Abstract

Federal agencies including the National Institutes of Health (NIH), Department of Defense (DoD) Congressionally Directed Medical Research Program (CDMRP) Spinal Cord Injury Research Program (SCIRP), and Department of Veterans Affairs (VA) provide the majority of funding for spinal cord injury (SCI) research in the United States. However, systematic evaluation of how funding is distributed across research areas, therapeutic approaches, and translational stages has been limited. To understand the distribution of funds, we curated and classified 1,589 federally funded SCI research awards from the NIH (2008–2023), the CDMRP SCIRP (2009–2023), and the VA (2017–2025). Each award was annotated based on the biological system or problem studied, the therapeutic intervention or approach utilized, and its placement along the translational continuum. Our analysis revealed that the NIH predominantly supports basic and early stage translational research, especially in areas of SCI pathology, regeneration, and motor functional recovery. In contrast, the CDMRP funding is more concentrated on applied and clinical research, particularly in the areas of pain, bladder function, and neuromodulatory device development. The VA predominantly invests in rehabilitation-focused studies and interventions aimed at improving musculoskeletal and functional health outcomes. While the complementary missions of these agencies collectively support a diverse SCI research ecosystem, we identified critical gaps in funding for high-priority areas such as bowel/gastrointestinal health, cardiovascular function, and mental health. Furthermore, the recent discontinuation of the CDMRP SCIRP and proposed NIH budget reductions are projected to lead to an approximate 50% decline in federal SCI research funding by 2026—posing a substantial risk to the field’s progress and threatening the stability of this ecosystem. These findings underscore the urgent need for coordinated, data-driven funding strategies that align more closely with the needs and priorities of the SCI community. To that end, we propose the development of a publicly accessible “living dashboard” to enhance transparency, foster interdisciplinary collaboration, and guide strategic investment in SCI research moving forward.

## INTRODUCTION

Spinal cord injury (SCI) is a devastating condition that often results in permanent motor, sensory, and autonomic dysfunction, profoundly impacting the quality of life of affected individuals. Currently, SCI affects over 300,000 people in the United States, with about 18,000 new cases annually^1^. In the absence of effective treatments, those living with SCI face loss of independence, a lifetime of health complications, and substantial barriers to full societal participation. The socioeconomic burden of SCI is correspondingly high, both in direct medical costs and in indirect costs related to long-term disability, lost productivity, and underappreciated caregiver needs. In 1997, the total economic burden of SCI to the US healthcare system was estimated at $7.74 billion^2^, a number which has only grown in the ensuing three decades, although more recent data are not available. These statistics illustrate the urgent need for targeted research aimed at improving health outcomes and quality of life for individuals living with SCI.

In the U.S., the majority of SCI research is funded by federal agencies; the NIH, the DoD (CDMRP SCIRP), and the VA represent the three principal contributors. Together, these agencies fund a diverse range of research, from basic studies of spinal cord biology to translational studies and clinical trials. Importantly, the missions of these three agencies are distinct: the NIH aims “to seek fundamental knowledge about the nature and behavior of living systems and the application of that knowledge to enhance health, lengthen life, and reduce illness and disability” (https://www.nih.gov/about-nih/what-we-do/mission-goals); the CDMRP SCIRP portfolio seeks “to fund research and encourage multidisciplinary collaborations for the development and translation of more effective strategies to improve the health and well-being of Service members, Veterans, and other individuals with spinal cord injury” (https://cdmrp.health.mil/scirp/default); whereas the VA Office of Research and Development describes its mission “to improve Veterans’ health and well-being via basic, translational, clinical, health services, and rehabilitative research”. These complementary goals establish the three federal agencies as three pillars upholding the SCI research ecosystem in the U.S.

In addition to federal agencies, private foundations such as the Craig H. Neilsen Foundation (CHNF) also contribute substantially (on the order of $20M annually; https://chnfoundation.org/#dollars-given), but federal investments remain the primary engine driving the national SCI research agenda. However, aside from the wealth of federal research funding data available from public databases such as the NIH Research Portfolio Online Reporting Tools Expenditures and Results (RePORTER), the Defense Technical Information Center (DTIC; https://dtic.dimensions.ai/discover/grant), and USAspending.gov, there does not yet exist a centralized, user-friendly system that allows for a “bird’s eye view” of how SCI research funding is distributed across scientific domains, therapeutic approaches, and stages of translational readiness. This lack of transparency makes it difficult for stakeholders to identify areas of momentum, redundancy, or unmet needs. Critically, while prior reports have evaluated the landscape of SCI clinical trials^3,4^, a comprehensive, systematic analysis of the federal SCI research funding ecosystem has not been conducted.

To address this critical knowledge gap, we curated and analyzed 1,589 federally funded SCI research awards spanning 2008 to 2025 within the NIH, CDMRP, and VA funding portfolios. Each award was categorized using a standardized classification schema developed with input from program officers, researchers, and individuals with lived SCI experience. Awards were categorized according to three axes: (1) the biological system or problem studied, (2) the therapeutic approach or intervention utilized, and (3) the stage of translational readiness. This structured analysis provides new insights into federal funding trends, agency-specific priorities, and gaps in research that may hinder scientific and clinical progress. Our findings also serve as a foundation for the development of a centralized, publicly accessible SCI research funding dashboard designed to inform strategic planning and support a more efficient, collaborative research ecosystem.

## METHODS

### Composition and contributions of the investigating team

The concept for this project was developed through discussion among members of the SCI advocacy organization Unite2Fight Paralysis (C. Barr, I. Burkhart, P. Nowell, A. G. Rabchevsky, and M. Rodreick) and SCI researchers (J. N. Dulin, A. G. Rabchevsky, and A. Torres-Espín). The data classification team was composed of individuals with lived SCI experience (C. Barr and J. Biundo), investigators in the SCI research field (J. N. Dulin, B. Kondiles, and D. A. McCreedy), and trainees (L. Abu, T. A. Barretto, J. Biundo, A. Buxton, S. Duenwald, A. Evans, T. N. Friedman, I. Gadaleta, S. Ganesh, T. Gillespie, B. Gottschall, P. Green, M. Leal-Garcia, G. Lee, L. Liu, R. N. Malik, E. J. Nava, M. R. Pacheco, C. Sorani, V. Tandon, H. Thomas, K. Thomas, A. V. Tran, and K. Vo). Team leaders (A. Buxton, J. N. Dulin, T. Gillespie, B. Kondiles, M. Leal-Garcia, M. Pacheco, A. V. Tran, and K. Vo) supervised the award classification process and resolved discrepancies. Data analysis was performed by C. Barr, J. Barr, H. Blackmon, J. N. Dulin, and A. Torres-Espín.

### Data retrieval and quality control

The search strategy used in this systematic analysis is shown in **Figure 1**. We adhered to the PRISMA 2020 reporting guidelines for systematic reviews^5^. For this study, we curated funding data from the NIH, the CDMRP, and the VA, as these are the three primary U.S. federal agencies that fund SCI research.

**Figure 1.**
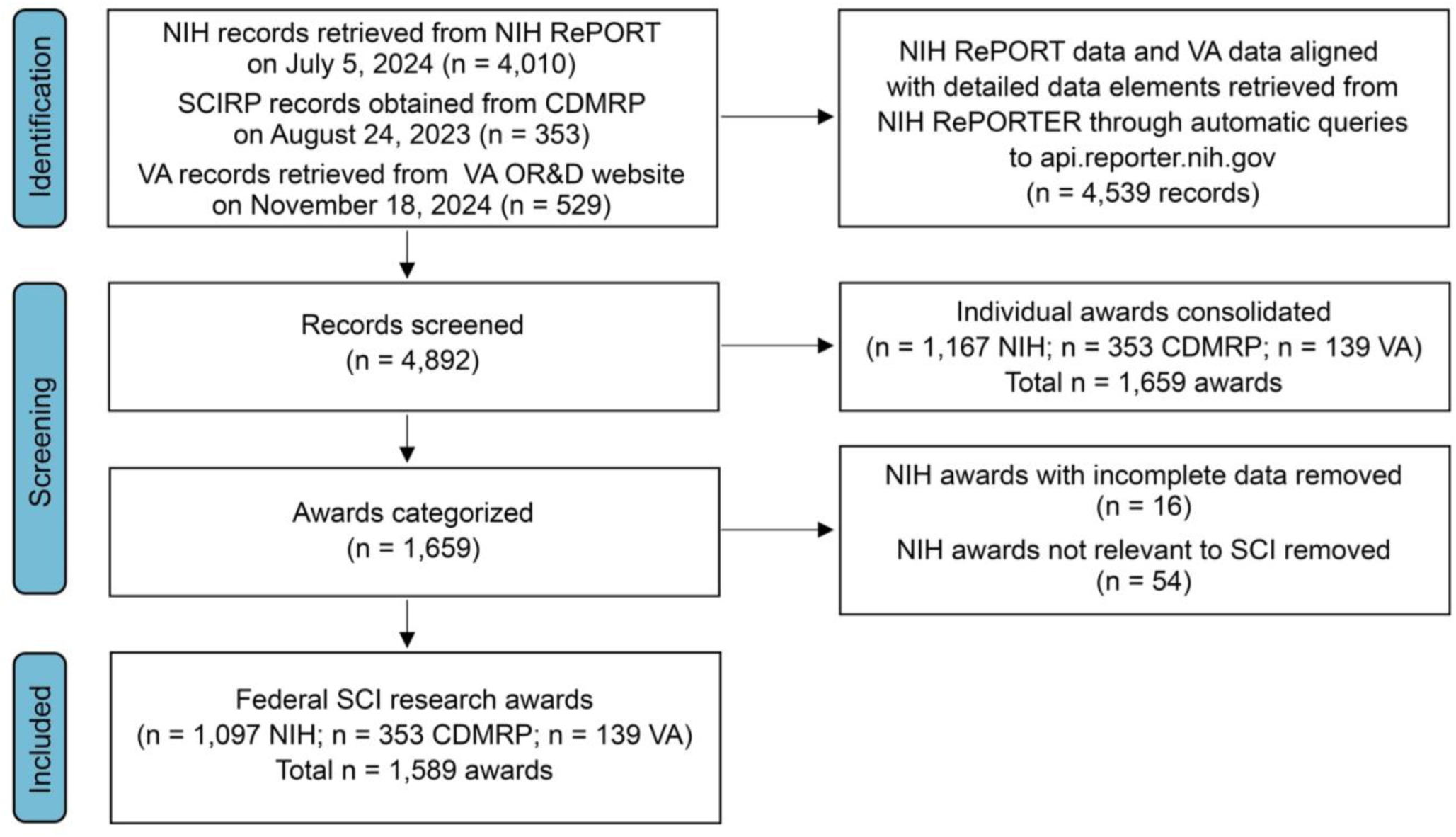
PRISMA flow diagram of the search strategy used in this study. CDMRP, Congressionally Directed Medical Research Program; DoD, Department of Defense; NIH, National Institutes of Health; OR&D, Office of Research & Development; SCI, spinal cord injury; VA, Department of Veterans Affairs.

#### National Institutes of Health data

To retrieve NIH data, awards within the “Spinal Cord Injury” research/disease area were downloaded for fiscal year (FY) 2008 – FY2023 on July 5, 2024 from the NIH Research Portfolio Online Reporting Tools (RePORT) Categorical Spending website (https://report.nih.gov/funding/categorical-spending#/; **Supplementary Table 1**). These were the only fiscal years for which NIH funding data were publicly available at the time of the search. We limited our search solely to grants that were listed in that portfolio; it is possible that other SCI-relevant awards exist outside of the RePORT “Spinal Cord Injury” category, but we did not expand our search to include these. Data were exported in *.XLSX format, and contained 4,010 records encompassing 1,167 unique projects (identified by their unique project serial numbers), with most awards spanning multiple fiscal years.

We next aligned the list of NIH awards retrieved from RePORT with more detailed data elements retrieved from NIH RePORTER (https://reporter.nih.gov/) through automatic queries to their Application Programming Interface (https://api.reporter.nih.gov/). During the alignment process, we curated the data in the following ways:

1. Consolidation of awards funded by multiple NIH Institutes and Centers (ICs): In the RePORT “Spinal Cord Injury” dataset, some grants appeared in multiple rows due to co-funding by different ICs; list of ICs is provided in **Supplementary Table 2**. For example, award 5K43TW010718-02 was listed with separate entries for the FIC ($50,306) and NINDS ($50,304). To avoid double-counting and overstating the number of unique grants, we consolidated these entries into a single row per award resulting in 93 awards funded by more than one IC. The total funding amount (inclusive of both direct and indirect costs) was calculated using NIH RePORTER data, and all contributing ICs were recorded in the designated Funding IC column.
2. Resolution of awards with incomplete data: For 10 awards, detailed information for individual subprojects was not available on NIH RePORTER, but dollar amounts for those subprojects were listed on RePORT (e.g., 1P01HD059751-01A1, which includes 5 subrecipients in FY2009 in the RePORT dataset). For those projects, we included the NIH RePORT dollar amount but based our classifications for all subprojects on the singular abstract available on NIH RePORTER. In addition, 15 awards listed on NIH RePORT (e.g., NTR12002001-1-0-1) were not found on NIH RePORTER. We were unable to classify these awards, so we excluded them from our dataset. For P01NS055976, a multi-PI project grant, some abstracts were not available on NIH RePORTER. Abstracts for these awards were provided by the Contact PI on the grant, Dr. John Houle (personal communications). One additional abstract did not contain useful information and was excluded from the dataset.
3. Resolution of awards with conflicting data: For 126 awards, we identified discrepancies in the dollar amounts listed on NIH RePORT (**Supp. Table 1**) and the dollar values listed on NIH RePORTER. For example, for 2R24HD050821-06, RePORT lists $100,000 to NIBIB, $626,707 to NICHD, and $200,000 to NINDS, for a total of $926,707 in FY2010. However, in NIH RePORTER the costs are listed as $1 to NIBIB, $21,944 to NICHD, and $1 to NINDS, for a total of $21,946 in FY2010. Together, these 126 awards resulted in a net discrepancy of $9,933,064 across FY2008-FY2023 that is listed by NIH RePORT but is not accounted for in NIH RePORTER. In these cases, we included only the RePORTER data in our analyses.

The complete dataset of NIH SCI awards is provided in **Supplementary Table 3**, with our award classifications in columns AK-AU. The list of excluded NIH awards, valued at an amount of $55.7M across FY2008-2023, is provided in **Supplementary Table 4**. A visual comparison of our data versus the raw RePORT data is shown in **Supplementary Figure 1**. On average, the mean absolute difference of our data versus the raw RePORT data is $5.47M, with a mean percent difference of 6.1% and a Pearson correlation coefficient of 0.90. Thus, our curated dataset trends very closely with RePORT data with a consistent negative bias due to the excluded awards and inclusion of RePORT (not RePORTER) data.

#### Department of Defense CDMRP data

CDMRP SCIRP data is publicly available in the DTIC database (https://dtic.dimensions.ai/discover/grant?search_mode=content&search_text=SCIRP&search_type=kws&search_field=full_search&or_facet_funder=grid.496791.4). A file containing 353 unique awards funded through the CDMRP SCIRP from FY2009-FY2023 was obtained from SCIRP program staff and included award information and associated research classifications data used internally for portfolio analysis (which are not available on DTIC) on August 24, 2023. CDMRP projects are not posted on NIH RePORTER, so more detailed data elements about these awards (e.g., annual direct and indirect costs, annual costs, institution, co-investigators), are not publicly available. The complete CDMRP dataset is provided in **Supplementary Table 5** with our modified award classifications in columns L-O.

#### Department of Veterans Affairs data

To retrieve the VA data, funded research projects for FY2017-FY2025 were obtained from the VA Office of Research & Development website (https://www.research.va.gov/about/funded_research/default.cfm) on November 18, 2024. These were the only fiscal years for which VA funding data were publicly available at the time of the search. Because the VA data comprised many awards spanning multiple programs, relevance of awards to spinal cord injury was determined by a team of 3 investigators, and only SCI-related grants were included in subsequent analysis. We obtained a total of 529 records encompassing 139 unique awards. Detailed data elements for these awards were retrieved from NIH RePORTER. The complete VA dataset is provided in **Supplementary Table 6**, with our award classifications in columns AC-AG. Note that the VA does not pay indirect costs, so all costs listed for VA awards are direct costs.

### Classification schema

The goal of data classification (categorization) was to provide descriptive information about individual awards according to metrics relevant to stakeholder groups including persons with lived experience, research investigators, clinicians, and federal and industry representatives. A three-axis classification system developed by CDMRP, NIH, VA, and CHNF staff was shared with the investigating team and used as a basis for this analysis. Classification categories were refined through group discussion among the classification team. This schema is provided in **Table 1**. Classification categories were developed and refined through group discussion among the classification team. The schema was designed to provide information about individual awards across three distinct axes: 1) System/Problem (“Why is this study being done, or what problem is being addressed?”), 2) Approach/Intervention (“What is the approach to be taken or the therapeutic intervention to be utilized?”), and 3) Readiness (“Where does this study lie on the spectrum from basic research to clinical research or treatment?”; **Figure 2**). Detailed information about the three-axis classification schema is provided in the **Supplementary Information** document.

**Table 1.** Classification schema used in this study. System/Problem, Approach/Intervention, and Readiness categories are included on separate tabs.

**Figure 2.**
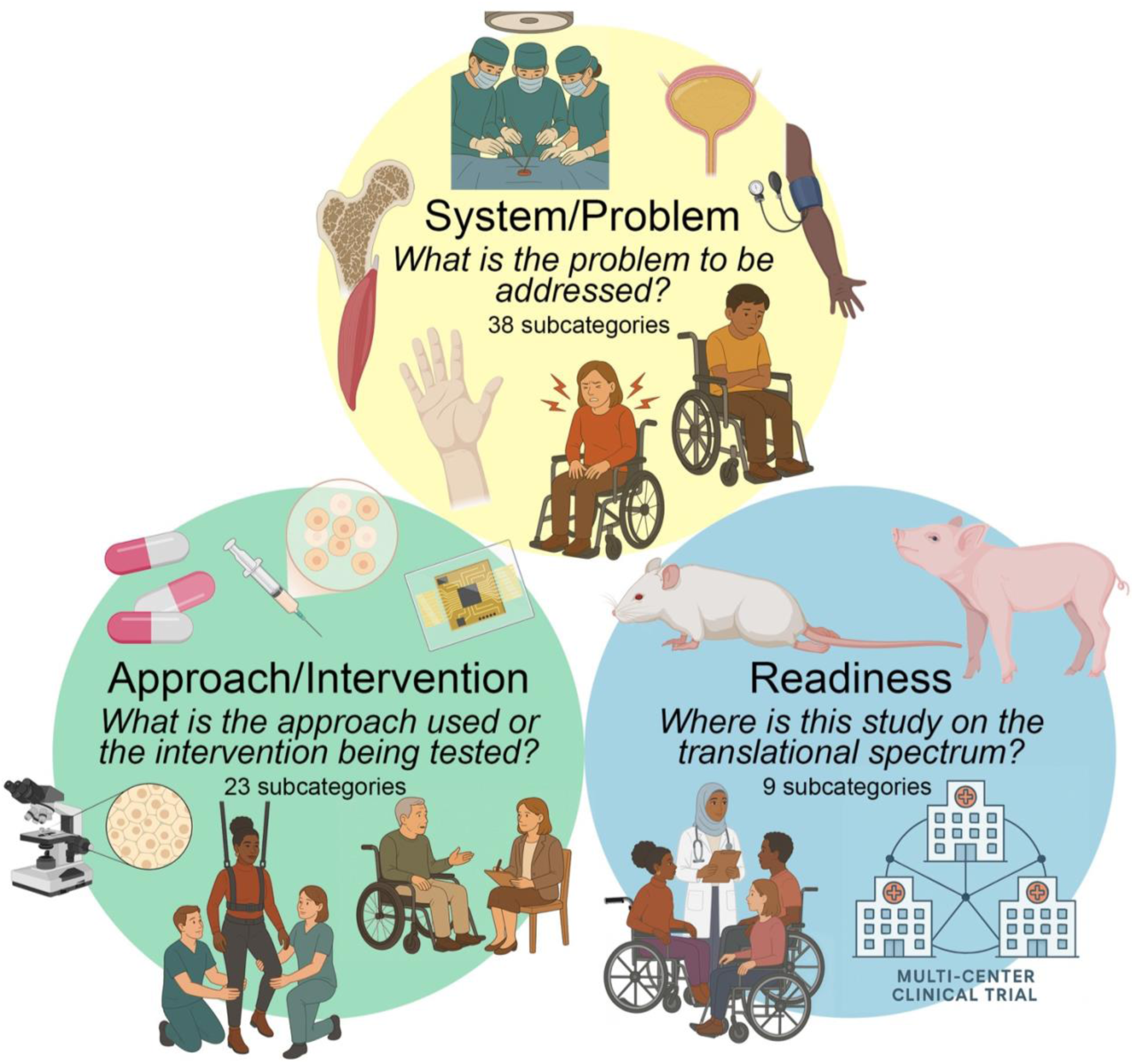
Graphical summary of the study classification schema. Federal grants were classified according to three axes: System/Problem, Approach/Intervention, and Readiness. Examples of specific subcategories are represented. Illustrations were created with Biorender and ChatGPT 4o.

### Data classification

Each award was reviewed by at least three members of the classification team. The NIH and VA awards were classified solely based on information provided in the project abstract, with any discrepancies among reviewers resolved through discussion with team leaders. CDMRP awards were provided by SCIRP program officers with preliminary classifications, which we then mapped to our standardized classification schema. If an abstract lacked sufficient detail to assign a classification in one or more of the three axes, the award was labeled “Unable to Classify” for that axis. All awards were assigned to a single primary subcategory within each axis. For research projects involving multiple approaches or therapeutic interventions, additional (secondary) descriptors were recorded in separate columns. These are referred to as “combinatorial” studies. Data related to combinatorial studies are presented in Figure 8; all other figures reflect analysis based solely on the primary classifications.

NIH awards determined to be not relevant to SCI were excluded from the dataset. These excluded awards did not involve spinal cord injury models or human subjects; did not focus on the development of tools or technologies applicable to SCI research or diagnosis; did not examine physiological systems in ways that would inform SCI; or otherwise lacked a clear connection to the field. In some cases, we applied expert judgment to assess relevance. For example, studies using fMRI to evaluate the effectiveness of body weight-supported treadmill training in stroke patients (e.g., NIH P41RR013642) were considered relevant to SCI, given the common use of this intervention in SCI rehabilitation. We also included awards that addressed nerve root trauma or developed imaging tools for spinal cord assessment in conditions such as multiple sclerosis or stroke.

Conversely, we excluded awards focused solely on vertebral or spinal column injury, multiple sclerosis pathology, peripheral nerve or optic nerve injury, stroke, traumatic brain injury, Alzheimer’s disease, and other neurological conditions that do not directly pertain to SCI. Representative examples of excluded awards include studies on Alzheimer’s disease, vertebral/spinal injuries, overactive bladder syndrome, and non-SCI-related pain. To maintain a focused dataset, these awards were omitted from further analysis. In total, we identified 70 NIH awards as not relevant to SCI (**Supp. Table 4**); the remaining 1,097 NIH awards were retained as relevant to SCI research (**Supp. Table 3**).

### Data analysis

Data were analyzed using Microsoft Excel, GraphPad Prism 10, and R Studio. To assess the relationships between funding distributions from different federal agencies, we performed pairwise correlation analyses using the XLMiner Analysis ToolPak in Microsoft Excel. This tool computes Pearson correlation coefficients, which measure the strength and direction of linear associations between pairs of continuous variables. Correlation values range from –1 to +1, with values closer to ±1 indicating stronger linear relationships. All data were first standardized where appropriate to account for differences in scale across funding agencies. Chi-Square tests with Bonferroni corrections were performed to determine whether allocation of funding among categories differed significantly among funding agencies. p-values < 0.05 were considered to be statistically significant. Detailed information about network analysis of combinatorial awards is provided in the **Supplementary Information** document.

## RESULTS

### Overview of federal funding

Our final dataset comprised 1,097 NIH awards, 353 CDMRP awards, and 139 VA awards, totalling 1,589 federally funded SCI research awards (**Table 2**). Annual expenditures, including both direct and indirect costs, are illustrated for each agency in **Figure 3**. Between 2008 and 2023, the NIH allocated approximately $67–$100 million annually to SCI research, with a median yearly investment of $77 million, for a total amount of $1.33B. From 2009 to 2023, the CDMRP contributed between $7–$37 million per year, with a median of $27 million, for a total amount of $389M. Since 2017, the VA has consistently invested between $9–$15 million annually, with a median of $12 million, for a total amount of $111M. (For comparison, the CHNF has awarded $422M across 2003-2024; https://chnfoundation.org/#dollars-given). The blue shaded region in **Figure 3** highlights the period from 2017 to 2023, during which expenditure data are available for all three federal agencies. Overall, these data indicate that federal funding for SCI research has remained relatively stable since 2019, with total expenditures equaling $139 million in 2023.

**Figure 3.**
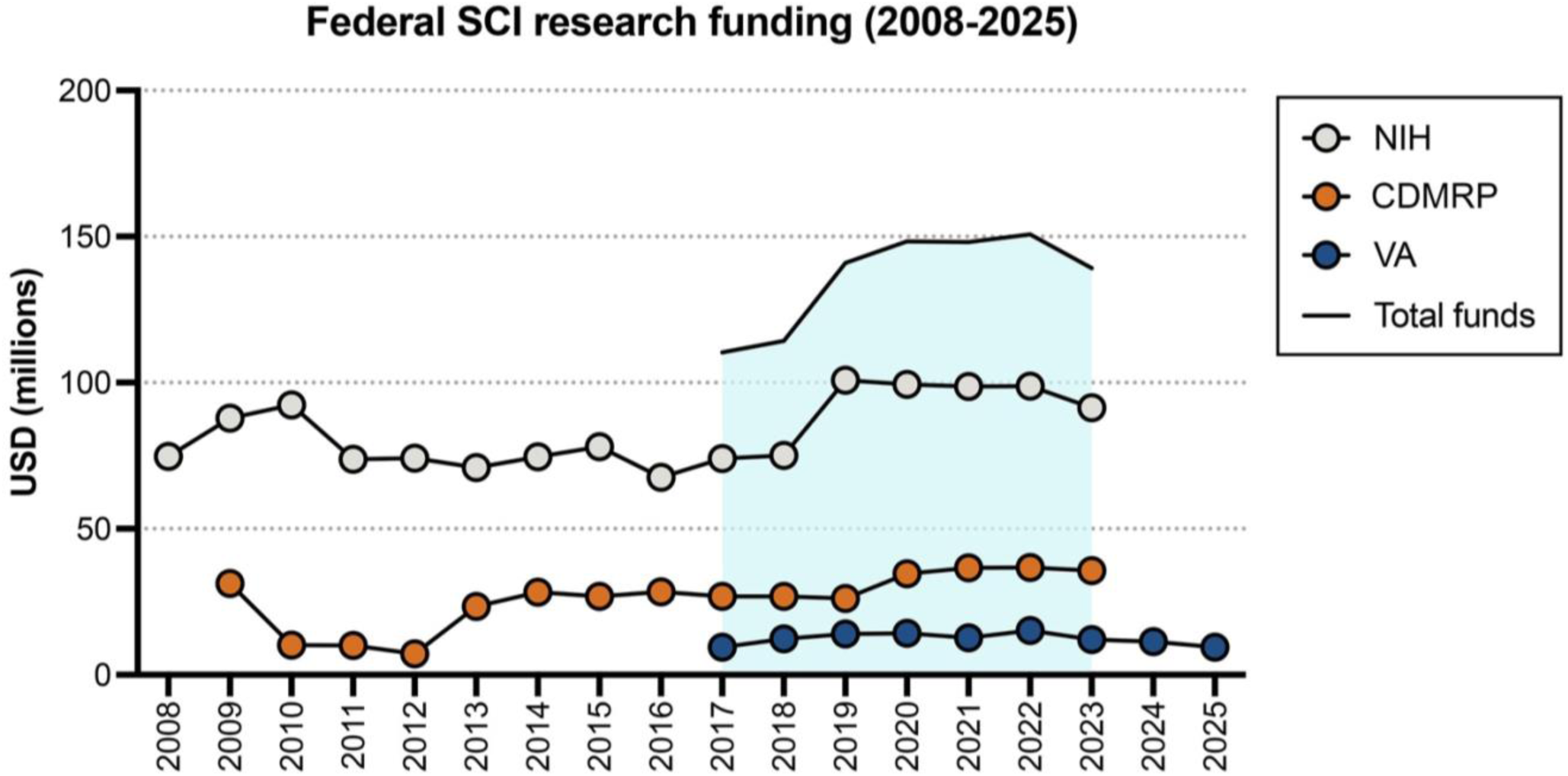
Federal spinal cord injury research funding from 2008-2025. Blue shaded area spans the years during which expenditure data are available for all three agencies (2017-2023). Data represent combined direct and indirect costs.

**Table 2.** Federal SCI research awards analyzed in this study. NIH, CDMRP, and VA awards are included on separate tabs.

Given that the NIH comprises 27 Institutes and Centers, along with other entities such as the Office of the Director (https://www.nih.gov/institutes-nih/list-institutes-centers; **Supp. Table 2**), we analyzed the distribution of SCI research funding across individual NIH components. As shown in **Supplementary Figure 2a**, the majority of NIH SCI research funding (66.1%) is awarded by the National Institute of Neurological Disorders and Stroke (NINDS). Other contributing institutes include the National Heart, Lung, and Blood Institute (NHLBI; 9.05%), the National Institute of Diabetes and Digestive and Kidney Diseases (NIDDK; 4.82%), and the Eunice Kennedy Shriver National Institute of Child Health and Human Development [NICHD; 3.49%, which houses the National Center for Medical Rehabilitation Research (NCMRR)], among others. NIH SCI awards span a range of funding mechanisms, with the largest proportion allocated to R-series (research) awards (75.8%), followed by U-series (cooperative agreements; 15.8%) and P-series (program project) awards (3.84%) (**Supp.** Fig. 2b). Funding categories representing less than 1% of the NIH SCI portfolio, such as F-series (fellowships; 0.75%) and T-series (institutional training grants; 0.046%), are not displayed in the figure. Across the NIH SCI research portfolio, 71.6% of funds are allocated to direct costs and 28.4% to indirect costs; comparable cost breakdowns were not available for CDMRP and VA awards. **Supplementary Figure 2**c highlights the 15 most highly funded NIH activity codes, with R01 awards accounting for 58.4% of total NIH SCI research funding.

Federal SCI research funding is awarded to many institutes across the U.S. (**Fig. 4**). For the time period of 2017- 2023, we found that the top ten states receiving SCI research funding include California ($140M), Pennsylvania ($92.3M), Ohio ($89.5M), New York ($63.2M), Florida ($59.2M), Kentucky ($52.3M), Texas ($48.4M), District of Columbia ($44.5M), Massachusetts ($41.9M), and Washington ($33.7M). In addition to the U.S., 2.7% of all federal funds during this time period were awarded to foreign entities including Canada, Australia, and others. A breakdown of funds awarded to state and international institutions by each agency is provided in **Supp.** Fig. 3.

**Figure 4.**
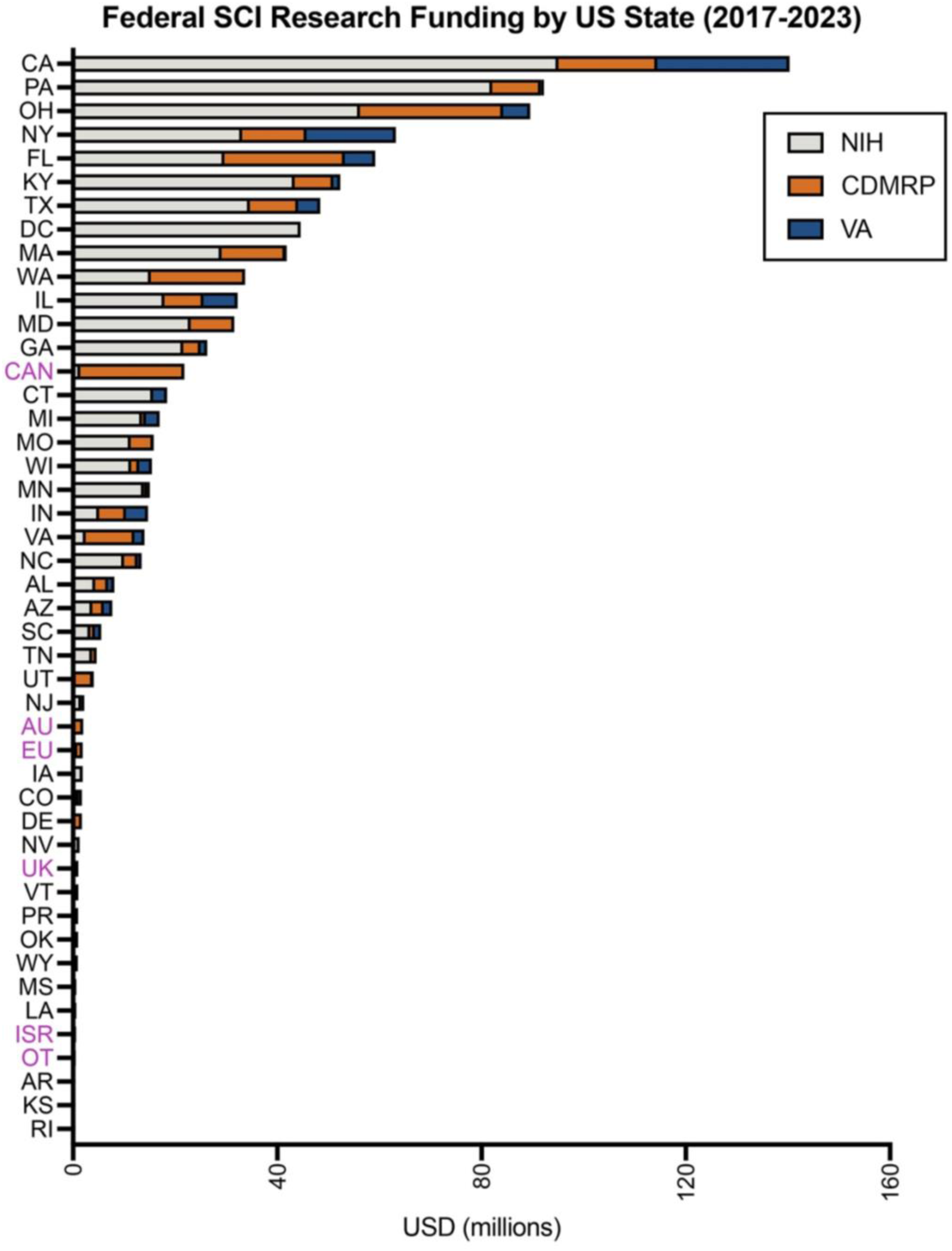
Spinal cord injury research funding by U.S. state. Combined federal funding for VA, DoD, and NIH for individual U.S. states for fiscal years 2008-2023. U.S. states that did not receive any federal SCI research funds during this time period are not shown. Non-U.S. entities are labeled in magenta: AU, Australia; CAN, Canada; EU, European Union; ISR, Israel; OT, other; UK, United Kingdom.

We also analyzed the top research institutions receiving federal SCI research funding from the NIH, CDMRP, and VA (**Supp.** Fig. 4). From FY2008-FY2023, Case Western Reserve University was the leading recipient of NIH funding, totaling $58.83M, followed closely by Drexel University with $58.80M. For CDMRP funding (2009– 2023), the University of British Columbia (a Canadian institution) ranked first with $23.4M, followed by the University of Miami Coral Gables ($22.3M) and Case Western Reserve University ($20.4M). Within the VA portfolio (2017–2025), the top recipients were the VA San Diego Healthcare System ($27.2M), the James J. Peters VA Medical Center ($18.8M), and the Edward Hines Jr. VA Hospital ($7.92M). These results are not surprising, as these institutions are among the most prominent and active spinal cord injury research centers in North America, with long-standing reputations in the field^6^.

In addition to institutional funding, we examined the distribution of awards across individual investigators from FY2008-FY2023 (**Supp.** Fig. 5). Among CDMRP-funded investigators, 74.5% received only one award, 19.3% received two, and just 6.2% of investigators received three or more awards. The NIH award distribution was highly right-skewed: more than half of NIH-funded investigators received three or fewer awards, with nearly 25% holding only one. A Shapiro-Wilk test confirmed a significant deviation from normality in the distribution (p < 0.001). In five cases, a single investigator received 30 or more NIH awards, with the highest individual total reaching 41 (this person was the contact PI on a large P grant with multiple subrecipients). In contrast, within the VA portfolio, the most common number of awards held per investigator was three (28.4%), a proportion greater than the combined total of investigators with one (5.68%), two (9.09%), or four (10.2%) awards. Collectively, these findings highlight distinctive differences in the distribution, and total amounts, of funding to SCI research across agencies, institutions, and investigators.

### Analysis of SCI research grants by system/problem

The general System/Problem categories used to classify federal SCI research awards are listed in **Table 1**. Between 2017 and 2023, 93.5% of awards fell into one of four primary categories: “SCI Motor/Sensory Function” (30.0%), “SCI Pathology & Repair” (27.7%), “SCI Secondary Health Conditions” (24.8%), and “SCI Adjacent” (11.0%) (**Fig. 5a**). The remaining 6.5% of awards were classified under “Acute SCI Management,” “Sociological,” “Infrastructure,” “Other SCI Studies,” or “Unable to Classify.” Differences in funding priorities among federal agencies are evident in the distribution of dollars across these categories. For example, 98.2% of the total funding for projects categorized as “SCI Adjacent” was provided by the NIH (**Fig. 5a**). In contrast, the CDMRP and VA allocated a greater proportion of their funds to “SCI Secondary Health Conditions” and “Motor/Sensory Function.” This divergence is further illustrated in **Figure 5b**, which compares the percentage of funding allocated to each category by individual agency. Specifically, the NIH allocated only 18.1% of its SCI research funding to “Secondary Health Conditions,” whereas the CDMRP and VA allocated 41.6% and 31.4%, respectively. To illustrate annual trends, **Figure 5c** presents a heat map showing the percentage of each agency’s total budget allocated to each category between 2017 and 2023. This visualization highlights agency-specific funding priorities—for instance, the CDMRP consistently allocated more of its annual budget to “Acute SCI Management” and “Sociological” studies compared to the NIH and VA, while the VA funded a greater proportion of “Infrastructure” grants in most years. A Chi-square analysis confirmed that the distribution of funding across categories significantly differs by agency, with p-values < 0.0001 after Bonferroni correction. Agency-specific breakdowns of these funding patterns are provided in **Supplementary Figure 6**.

**Figure 5.**
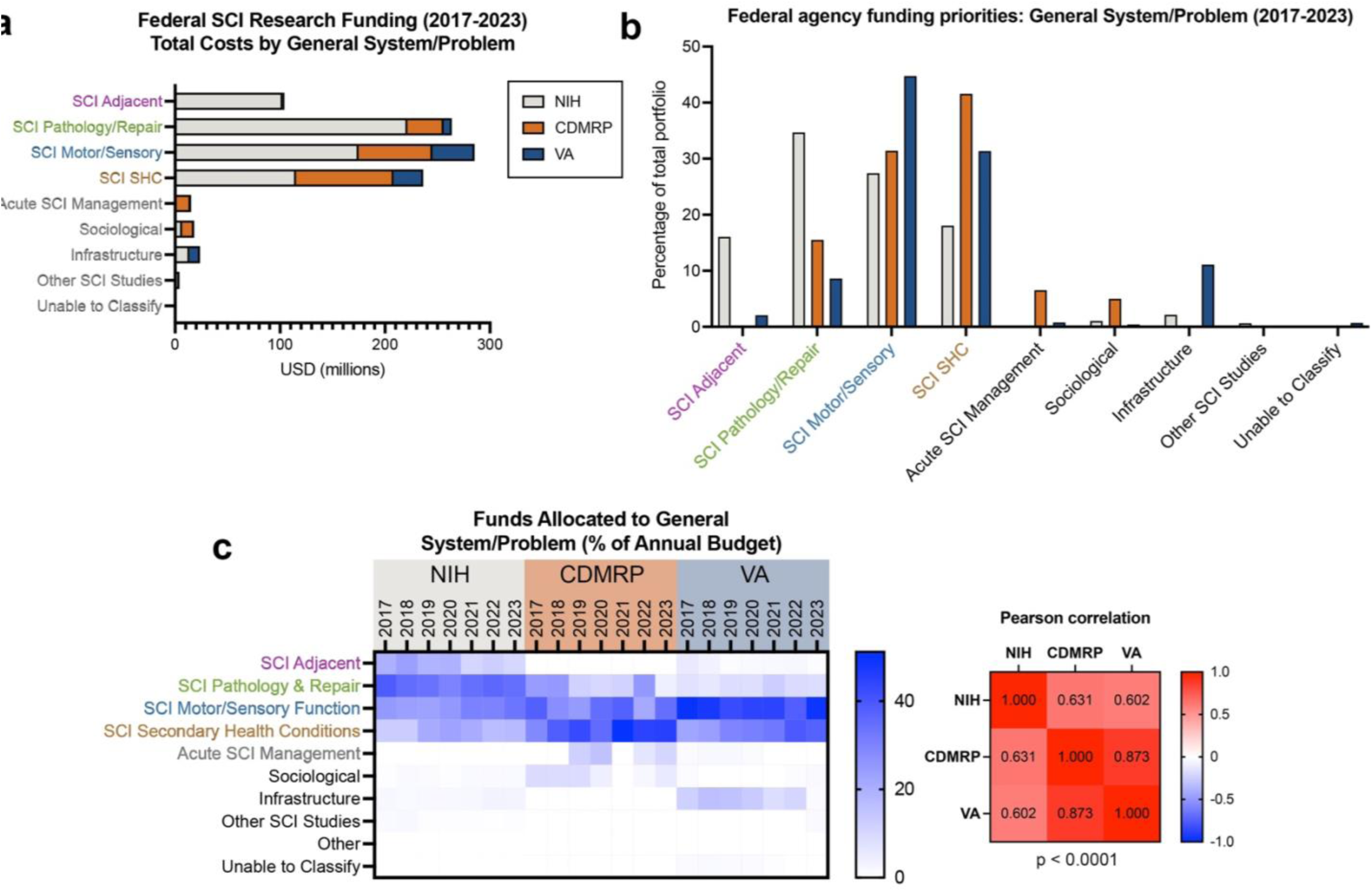
Federal SCI research funding categorized by general system/problem. (**a**) Breakdown of total federal SCI research funding from 2017-2023 by general system/problem category. The color coding of SCI Adjacent, SCI Pathology/Repair, SCI/Motor Sensory, and SCI Secondary Health Conditions (SHC) corresponds to the color coding in Figures 6, 11, and 12. (**b**) Proportions of each federal agency’s budget allocated to each general system/problem category; data are expressed as a proportion of 100% for each agency. (**c**) Heat map of federal funds allocated to each category for individual years 2017-2023; color scale reflects percent of annual budget. Each column totals 100%. Pearson correlation coefficients of funding priorities among the three agencies are shown to the right, with the results of Chi-square analysis showing a statistically significant difference in the distribution of funding across agencies (p < 0.0001).

In addition to the general System/Problem categories presented in Figure 5, we further classified awards into more specific subcategories to capture the targeted physiological or functional issues addressed by each project (**Table 1**). These detailed classifications include areas such as bladder function, breathing, and sleep, offering greater resolution into the specific problems investigated through federally funded SCI research. **Figure 6** displays the total funding, in millions of dollars, allocated to each specific system/problem subcategory by the NIH, CDMRP, and VA during the 2017–2023 period. This figure highlights notable differences in agency-level funding priorities. For example, the NIH invested $157M in studies focused on regeneration, plasticity, and repair, compared to $10.8M and $4.92M from the CDMRP and VA, respectively. These mechanistic regeneration/repair studies are typically earlier in development as they focus on assessing molecular or cellular outcomes rather than functional recovery. Certain subcategories received funding exclusively from a single agency; for instance, research on the intact spinal cord was funded solely by the NIH, while studies related to Acute SCI Management were supported only by the CDMRP ($14.6M) and the VA ($0.702M). These findings underscore the distinct programmatic missions and research emphases of each funding agency. **Figure 6b** displays changes in funding priorities over time. While the NIH and VA portfolios appear relatively consistent, changes in funding priorities are evident in the CDMRP dataset; for example, areas including breathing, integument, psychological/psychosocial, and AD/autonomic function receive funding in some years but not others, and even the frequency of funding for hindlimb and forelimb motor studies varies from year to year. This is likely due to differences in the review and funding recommendation processes among the difference agencies, as CDMRP does not employ a payline and applications must address specific programmatic goals that are re-evaluated on an annual basis and communicated in the funding opportunity announcements. A Chi-square analysis confirmed that the distribution of funding across categories significantly differs by agency, with p-values < 0.0001 after Bonferroni correction.

**Figure 6.**
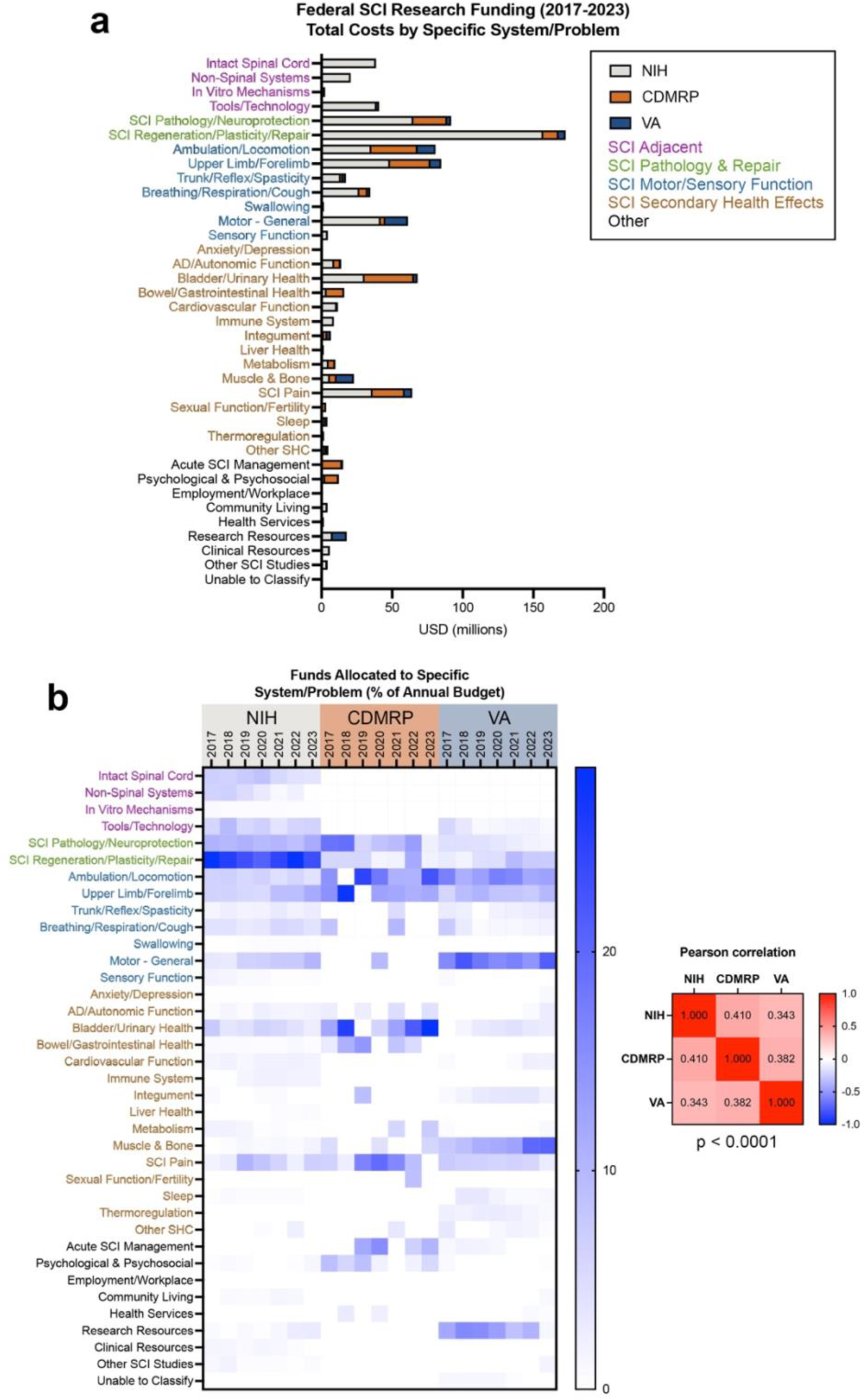
Federal SCI research funding categorized by specific system/problem. (**a**) Breakdown of total federal SCI research funding from 2017-2023 by specific system/problem category. Specific categories are color- coded according to the general system/problem category. (**b**) Heat map of federal funds allocated to each specific system/problem category for individual years 2017-2023; color scale reflects percent of annual budget. Each column totals 100%. Pearson correlation coefficients of funding priorities among the three agencies are shown to the right, with the results of Chi-square analysis showing a statistically significant difference in the distribution of funding across agencies (p < 0.0001).

When represented as percentages of each agency’s total SCI research budget, distinct differences in federal funding priorities become more evident (**Supp.** Fig. 7a). A significant portion of the VA portfolio is directed toward secondary health conditions, including muscle and bone health (13.6%), research resources (11.1%), lower extremity motor function (14.1%), and general motor function (17.7%). In contrast, CDMRP funding emphasizes areas such as bladder health (15.7%) and SCI-related pain (10.2%), consistent with the priorities outlined by the SCIRP (https://cdmrp.health.mil/scirp/default). The NIH, as previously described, allocates a substantial portion of its SCI research funding to regeneration (24.6%) and SCI pathology/neuroprotection (10.2%), while comparatively fewer funds are directed toward secondary health conditions such as bladder health (4.78%) and pain (5.67%). Some areas, including autonomic dysreflexia, receive relatively minimal attention across all agencies, accounting for less than 1.5% of total federal SCI research funding. Detailed agency-specific breakdowns of specific system/problem subcategories are provided in **Supplementary** Figures 7 and 8.

### Analysis of SCI research grants by approach/intervention

The Approach/Intervention categories used to classify federal SCI awards are listed in **Table 1**. These categories were broadly grouped into three overarching domains: “Discovery/Tools,” which includes studies that do not involve therapeutic interventions; “Therapeutic Interventions,” which includes studies employing one or more therapeutic approaches; and “Community/Resources,” which encompasses awards supporting psychosocial programs, community engagement, or research infrastructure. **Figure 7a** presents the total funding, in millions of dollars, allocated to each specific Approach/Intervention category by the NIH, CDMRP, and VA from 2017 to 2023. Among all categories, “In vivo approaches”—representing animal studies that do not test therapeutic interventions—received the highest level of federal investment, totaling $191M during the study period. Strikingly, grants in this category were almost entirely (97.6%) funded by NIH. Substantial funding was also directed toward invasive devices ($155M), noninvasive devices ($96.1M), drug interventions ($151M), and biological approaches, including biomaterials ($14.2M), cell therapies ($50.2M), and other biologics ($72.6M).

**Figure 7.**
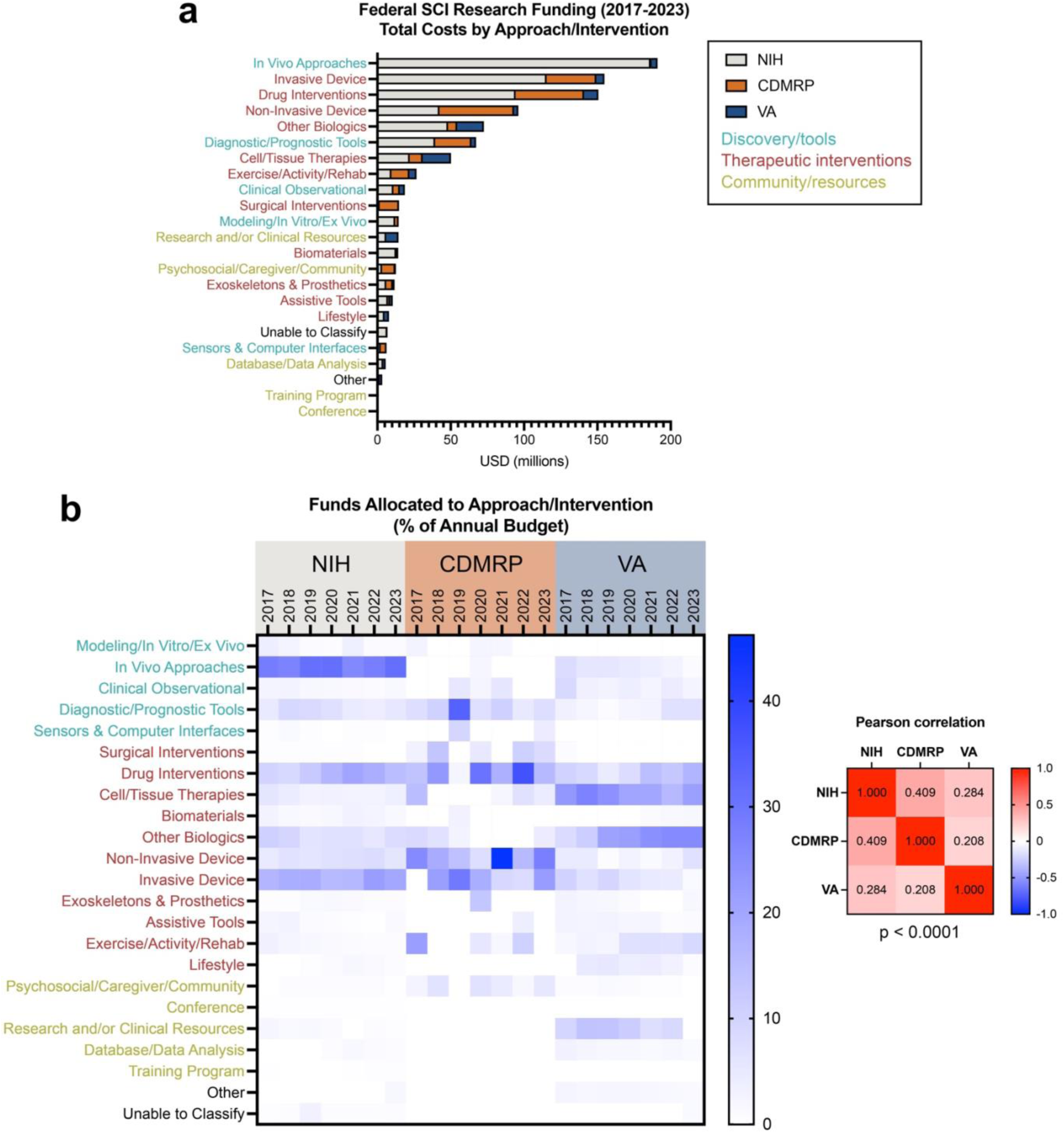
Federal SCI research funding categorized by approach/intervention. (**a**) Breakdown of total federal SCI research funding from 2017-2023 by approach/intervention category, sorted by most- to least- funded. (**b**) Heat map of federal funds allocated to each approach/intervention category for individual years 2017- 2023; color scale reflects percent of annual budget. Each column totals 100%. Pearson correlation coefficients of funding priorities among the three agencies are shown to the right, with the results of Chi-square analysis showing a statistically significant difference in the distribution of funding across agencies (p < 0.0001).

**Supplementary Figure 9** displays the proportion of each agency’s budget allocated to the various Approach/Intervention categories. Notable differences in funding priorities emerged among agencies. For instance, “In vivo approaches” constituted 29.2% of the NIH’s SCI budget but only 0.31% and 4.42% of the CDMRP and VA budgets, respectively. Conversely, the VA invested a relatively large share of its SCI research portfolio in cell/tissue therapies (21.4%), compared to the NIH (3.45%) and CDMRP (3.96%). The CDMRP devoted a higher proportion of its budget to noninvasive devices (22.7%) than either the NIH (6.64%) or VA (3.27%). Chi-square analysis confirmed that these differences in budget allocation across categories and agencies were statistically significant (p < 0.0001, Bonferroni corrected). Temporal trends in funding distribution across Approach/Intervention categories and agencies are shown in **Figure 7b**. Consistent with the System/Problem analysis, relatively few changes are observed over time within the NIH and VA portfolios, which exhibit stable distributions across most categories. In contrast, the CDMRP portfolio shows greater variability over the study period, particularly in categories such as “Exercise/Training/Rehabilitation,” “Surgical Interventions,” and “Cell/Tissue Therapies.” Further detailed breakdowns of Approach/Intervention categories for each agency are provided in **Supplementary Figure 10**. Overall, these data show that while the NIH and VA maintain consistent investment patterns across intervention types, the CDMRP demonstrates more dynamic shifts in funding trends, potentially underscoring more investment in emerging therapeutic strategies and translational approaches in SCI research.

For simplicity, awards were analyzed based on their primary Approach/Intervention category. However, a subset of awards—particularly within the NIH portfolio—employed combinatorial approaches. Among the 1,097 NIH SCI research awards analyzed, 152 (13.9%) involved more than one intervention: 133 awards utilized two interventions, 16 utilized three, and 3 incorporated four distinct approaches (**Fig. 8a**). The frequency with which individual intervention types appear in these combinatorial awards is shown in **Figure 8b**. Network analysis of combinatorial SCI research awards revealed a highly structured pattern of intervention pairing ( **Fig. 8c**). The most frequently co-utilized approaches were cell/tissue therapies combined with other biologicals, reflecting the prominent role of cell-based strategies in preclinical SCI research. Other enriched combinations included device-based interventions with exercise/rehabilitation and, highlighting the translational emphasis on multimodal strategies. Overall, nine intervention pairs occurred more frequently than expected under the independence model (p < 0.05, Bonferroni corrected; **Table 3**), suggesting deliberate scientific and therapeutic rationales for these combinations. These findings underscore a growing trend toward integrative approaches in SCI research and point to specific areas where synergistic interventions are being prioritized in the federal funding landscape.

**Table 3.** Significantly enriched combinatorial intervention pairs in federally funded SCI research. Table displays the nine intervention pairs that were combined more frequently than expected under an independence model (Bonferroni-adjusted p < 0.05). Observed pairings reflect the number of awards in which each intervention pair co-occurred; expected pairings represent the modeled frequency based on the marginal prevalence of each intervention across all awards. p-values were calculated using a one-tailed Poisson test with Bonferroni correction for 47 comparisons.

**Figure 8.**
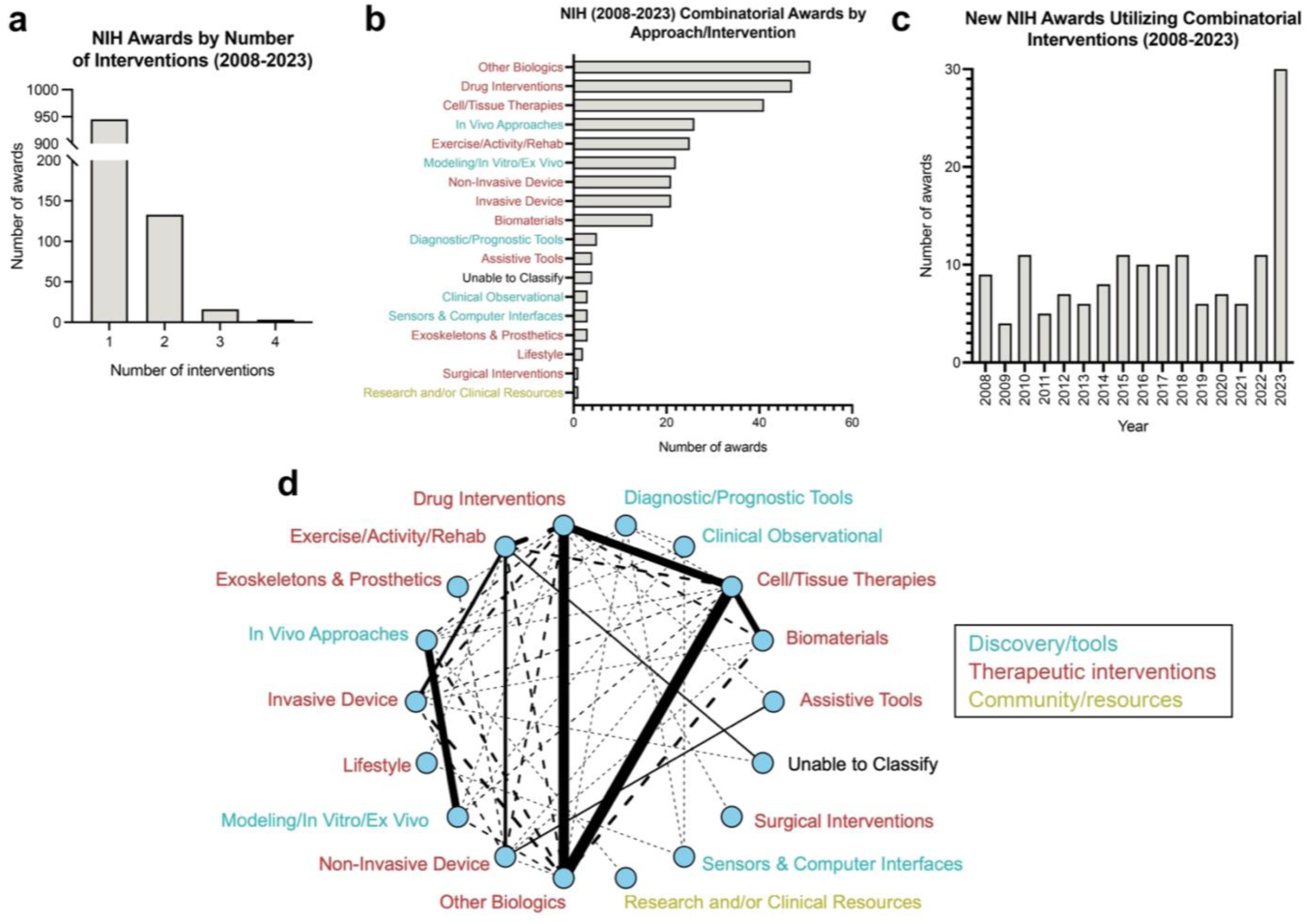
Analysis of NIH research awards utilizing combinatorial interventions. (**a**) Number of NIH SCI research awards according to the number of approach/interventions used. (**b**) The number of combinatorial awards utilizing each type of approach/intervention. (**c**) The number of new awards each year utilizing combinatorial interventions. (**d**) Network visualization of combinatorial intervention pairings. Each node represents a distinct therapeutic intervention category; edges represent observed co-occurrences of intervention pairs within individual awards. Edge width is scaled to the frequency of observed pairings. Solid edges indicate statistically significant enrichment (Bonferroni-adjusted p < 0.05). Dashed edges indicate non-significant pairings. Statistical significance was assessed using a one-tailed Poisson test under an independence model.

### Analysis of SCI research grants by readiness

We also analyzed the distribution of funding across distinct Readiness categories (**Table 1**), which reflect the translational stage of the research (**Fig. 9**). From 2017 to 2023, a total of $617M was allocated to *in vitro* and animal studies—including basic-basic, disease-related basic, and preclinical research—while $292M supported clinical studies (**Fig. 9a**). The proportion of each agency’s budget allocated to individual Readiness categories is shown in **Figure 9b**. One striking observation is that nearly all federal support for Phase I/II clinical trials and clinical feasibility studies originated from the CDMRP, which allocated 39.4% of its SCI research budget to this category. In comparison, the NIH and VA devoted only 0.63% and 0.00% of their budgets, respectively, to these early-phase clinical studies. Conversely, all basic-basic research awards were funded exclusively by the NIH, comprising 8.02% of its total SCI research portfolio. Despite these differences, all three agencies devoted substantial portions of their budgets to preclinical studies: 37.4% for the NIH, 31.3% for the CDMRP, and 53.1% for the VA. Temporal trends in budget allocation by Readiness category are shown in **Figure 9c**. These data illustrate the relative consistency of each agency’s investment patterns across translational stages over time. Notably, the VA has consistently allocated a portion of its budget to infrastructure, which is expected given that VA research is conducted exclusively within VA institutions and often requires internal support for clinical and research infrastructure development. Chi-square analysis confirmed that the distribution of funding across categories differs significantly among the three agencies (p < 0.0001). Detailed breakdowns for each agency’s full dataset are provided in **Supplementary Figure 11**.

**Figure 9.**
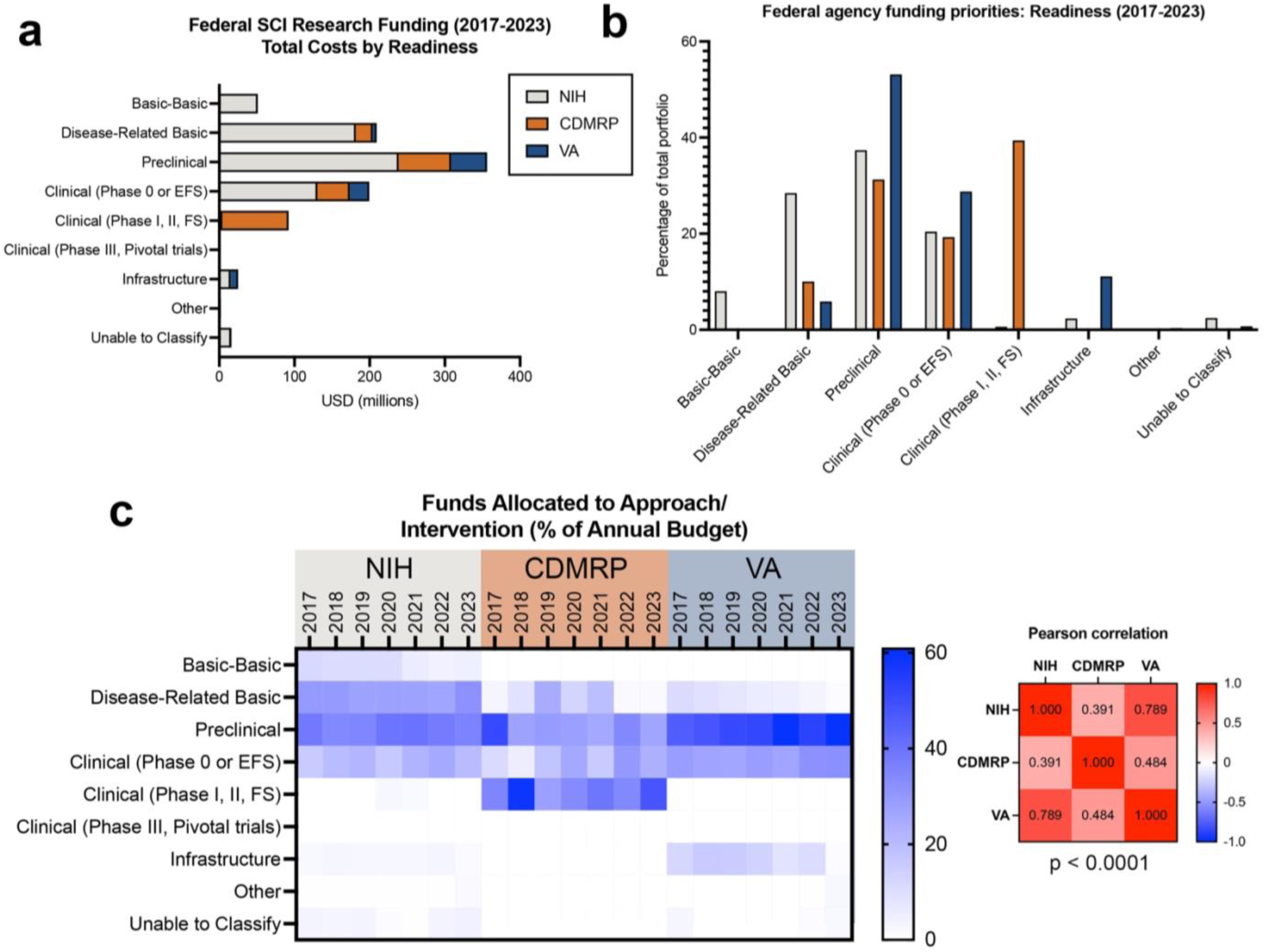
Federal SCI research funding categorized by readiness. (**a**) Breakdown of total federal SCI research funding from 2017-2023 by readiness category. (**b**) Proportions of each federal agency’s budget allocated to each readiness category; data are expressed as a proportion of 100% for each agency. (**c**) Heat map of federal funds allocated to each category for individual years 2017-2023; color scale reflects percent of annual budget. Each column totals 100%. Pearson correlation coefficients of funding priorities among the three agencies are shown to the right, with the results of Chi-square analysis showing a statistically significant difference in the distribution of funding across agencies (p < 0.0001).

### Intersectional analyses

Given the global trends observed in these datasets, we next sought to examine how categories of federal SCI research funding intersect with one another. As a first step, we analyzed the predominant Approach/Intervention types used within each Readiness category (**Fig. 10**). Among basic research awards, which include both basic-basic and disease-related basic studies, the majority of funding (70.7%) supported in vivo approaches (**Fig. 10a**). Additional investments were directed toward research tool development (12.7%), modeling/in vitro/ex vivo approaches (5.47%), and clinical observational studies (5.27%). In the preclinical readiness category, which includes animal studies involving therapeutic interventions, the largest proportion of funding supported drug-based interventions (34.0% of preclinical funding; **Fig. 10b**). Other frequently used preclinical approaches included invasive and noninvasive neuromodulatory devices (16.9% and 5.69%, respectively), as well as biological therapeutics, including cell/tissue therapies (13.6%), biomaterials (3.70%), and other biologics (16.9%). In clinical studies, more than half of the total funding was allocated to neuromodulatory device interventions; 28.8% for invasive devices and 25.6% for noninvasive devices (**Fig. 10c**). In contrast, only 9.19% of clinical research funding supported drug interventions, 7.63% supported tool development. This trend may be due to the high cost of device studies compared to studies utilizing other interventions. Interestingly, only 5.24% of clinical research funding was allocated to exercise and rehabilitation studies; this is particularly notable given prior findings that rehabilitation is the most frequently used intervention in SCI clinical trials^3^, suggesting either a disconnect between funding allocation and clinical practice trends, or a relatively lower cost of rehabilitation studies. **Figure 10d** provides a comparative summary of intervention-specific funding priorities across all readiness categories.

**Figure 10.**
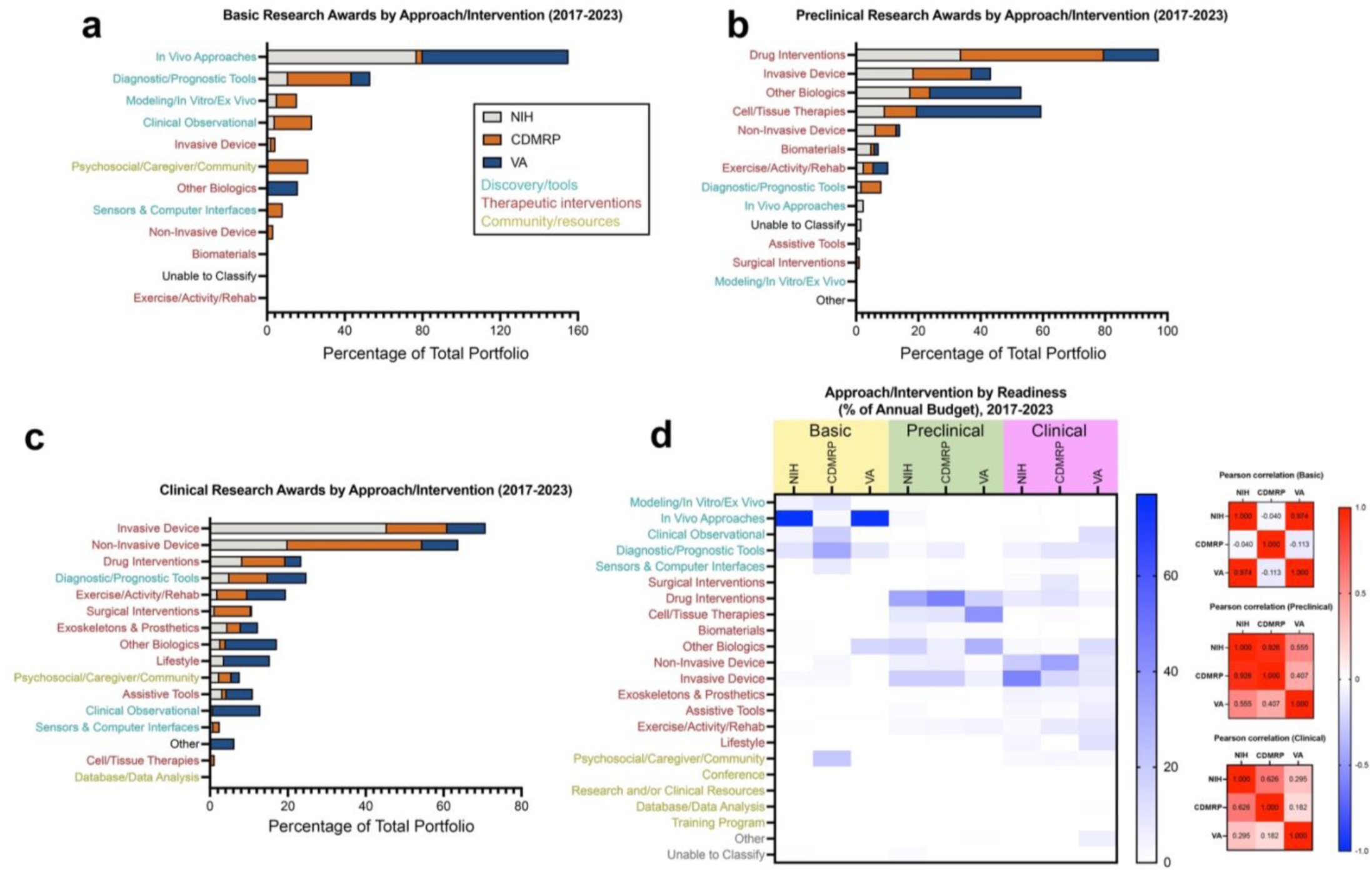
Intersectional analysis of readiness categories by approach/intervention used, 2017-2023. (**a**) Federal funding for basic research awards, including basic-basic and disease-related basic awards, expressed as a percentage of each agency’s total budget between 2017-2023 according to the approach/intervention used. (**b**) Federal funding for preclinical research awards expressed as a percentage of each agency’s total budget between 2017-2023 according to the approach/intervention used. (**c**) Federal funding for preclinical research awards, including Phase 0, Phase I/II, and Phase III/IV awards, expressed as a percentage of each agency’s total budget between 2017-2023 according to the approach/intervention used. Approach/intervention categories that received no funding during this time period are not shown for panels a-c. (**d**) Heat map of federal funds allocated to each approach/intervention category within each readiness bin; color scale reflects percent of annual budget. Each column totals 100%. Pearson correlation coefficients of funding priorities among the three agencies are shown to the right.

We also examined how SCI research awards at different translational stages (basic, preclinical, and clinical) are distributed across specific System/Problem categories. For basic research grants, the largest proportion of funding was directed toward studies of SCI regeneration/plasticity/repair (24.8%) and SCI pathology/neuroprotection (16.7%), followed by studies of the intact spinal cord (11.9%) and non-spinal systems (6.18%; **Fig. 11a**). Preclinical awards similarly prioritized SCI regeneration/plasticity/repair (29.0%) and SCI pathology/neuroprotection (10.7%), with additional significant investments in general motor function (10.1%) and bladder function (10.1%; **Fig. 11b**). As expected, clinical awards showed a shift in focus toward functional outcomes, with the largest proportion of funding supporting upper limb/forelimb function (19.3%), followed by ambulation/locomotion (17.0%), SCI pain (8.90%), and bladder/urinary health (7.92%; **Fig. 11c**). A detailed breakdown of funding allocations into System/Problem categories for each agency’s basic, preclinical, and clinical portfolios is provided in **Figure 11d**.

**Figure 11.**
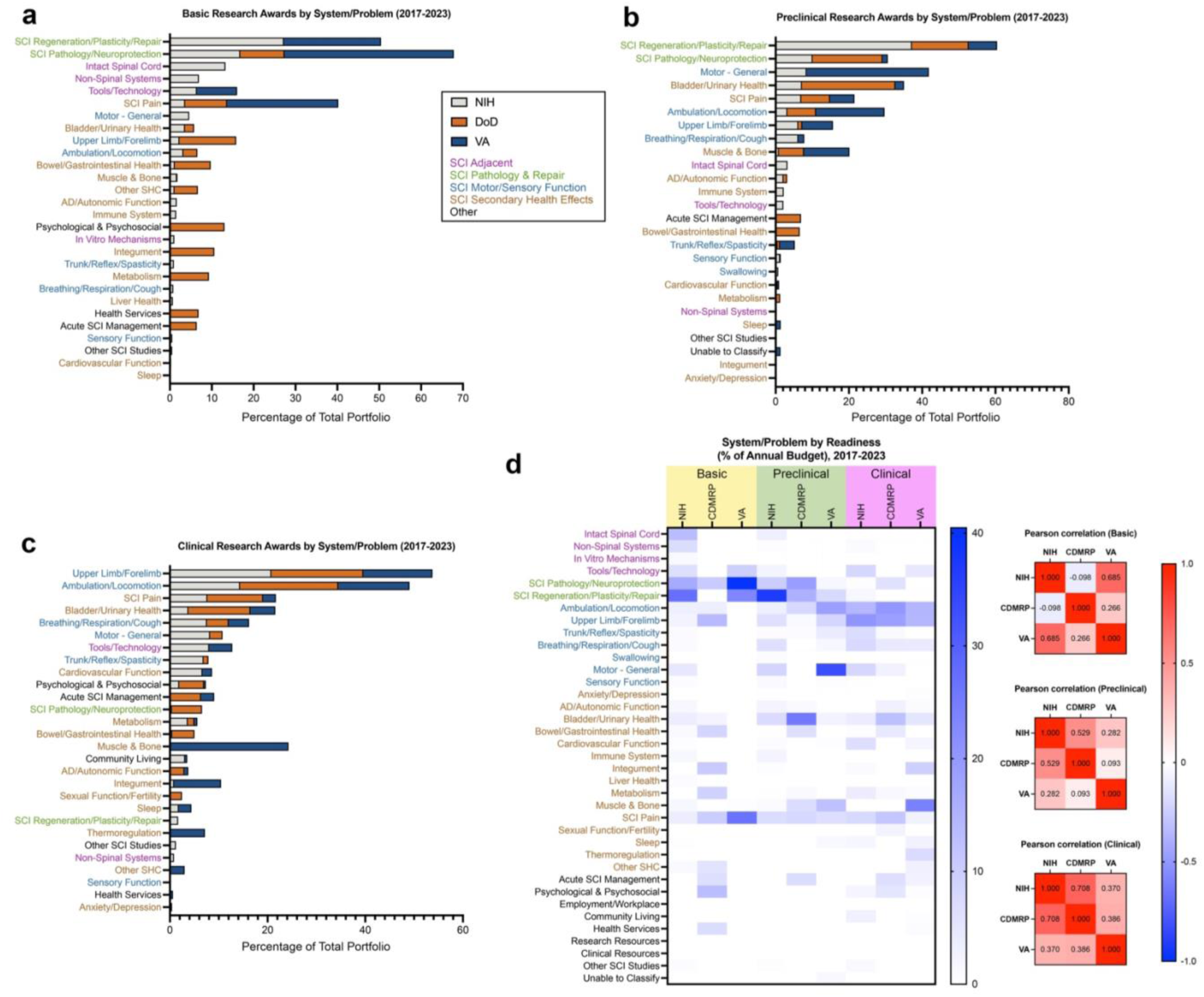
Intersectional analysis of readiness categories by system/problem studied, 2017-2023. (**a**) Federal funding for basic research awards, including basic-basic and disease-related basic awards, expressed as a percentage of each agency’s total budget between 2017-2023 according to the specific system/problem addressed. (**b**) Federal funding for preclinical research awards expressed as a percentage of each agency’s total budget between 2017-2023 according to the specific system/problem addressed. (**c**) Federal funding for clinical research awards, including Phase 0, Phase I/II, and Phase III/IV awards, expressed as a percentage of each agency’s total budget between 2017-2023 according to the specific system/problem addressed. Approach/intervention categories that received no funding during this time period are not shown for panels a-c. (**d**) Heat map of federal funds allocated to each system/problem category within each readiness bin; color scale reflects percent of annual budget. Each column totals 100%. Pearson correlation coefficients of funding priorities among the three agencies are shown to the right.

Finally, we evaluated how research funds for specific therapeutic interventions are distributed across System/Problem categories to better understand how different treatments are being developed to address specific aspects of SCI-associated pathologies. We focused on four major classes of interventions that receive a large proportion of federal funds: (1) biological interventions (cell/tissue therapies, biomaterials, and other biologics), (2) device-based interventions (both invasive and non-invasive), (3) drug interventions, and (4) exercise/activity/rehab. For awards utilizing biological interventions, 36.7% of funding supported SCI regeneration/plasticity/repair, 29.3% supported motor/sensory functional studies, and 8.5% supported bladder function/health (**Fig. 12a**). Within this category, the VA portfolio was primarily directed toward general motor function (39.7%), muscle and bone health (17.0%), and ambulation/locomotion (16.7%). In contrast, the NIH allocated 52.2% of its biological intervention funding to SCI regeneration/plasticity/repair and 12.9% to SCI pathology/neuroprotection, both of which categories do not assess functional outcomes. The CDMRP portfolio emphasized bladder function/health (27.1%), SCI regeneration/plasticity/repair (25.3%), and SCI pain (13.9%).

**Figure 12.**
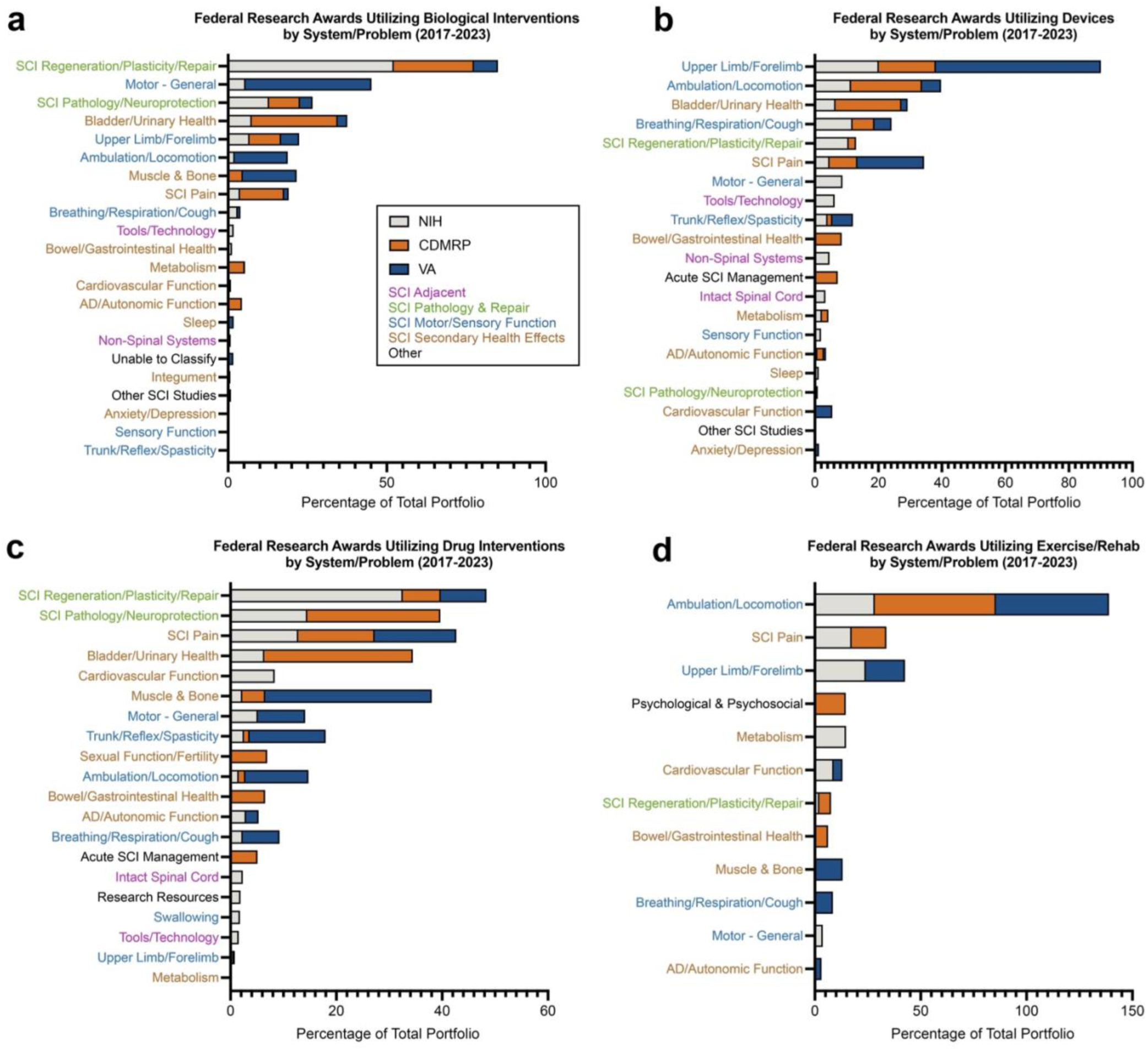
Intersectional analysis of selected therapeutic interventions by system/problem studied, 2017- 2023. (**a**) Federal funding for awards utilizing cell therapies, biomaterials, or other biologics, expressed as a percentage of each agency’s total budget between 2017-2023 according to the specific system/problem addressed. (**b**) Federal funding for awards utilizing invasive- or non-invasive devices, expressed as a percentage of each agency’s total budget between 2017-2023 according to the specific system/problem addressed. (c) Federal funding for awards utilizing drug interventions, expressed as a percentage of each agency’s total budget between 2017-2023 according to the specific system/problem addressed. (d) Federal funding for awards utilizing exercise, training, or rehabilitation, expressed as a percentage of each agency’s total budget between 2017 - 2023 according to the specific system/problem addressed.

For device-based interventions, the leading system/problem categories were upper limb/forelimb function (20.5% of total funds), ambulation/locomotion (14.9%), and bladder function/health (11.2%; **Fig. 12b**). Notably, the VA allocated over half (51.9%) of its device-related funding to upper limb/forelimb studies, followed by SCI pain (21.0%). NIH device studies were more evenly distributed, with funding directed toward upper limb/forelimb (20.2%), breathing/respiration/cough (11.9%), ambulation/locomotion (11.4%), and SCI regeneration/plasticity/repair (10.7%). The CDMRP prioritized ambulation/locomotion (22.4%), bladder function/health (20.8%), and upper limb/forelimb (18.0%).

Among awards utilizing drug-based interventions, the top system/problem categories were SCI regeneration/plasticity/repair (23.2%), SCI pathology/neuroprotection (16.9%), and SCI pain (13.5%; **Fig. 12c**). Within this category, the VA focused on muscle and bone health (31.5%) and SCI pain (15.4%). The CDMRP allocated over half of its drug-related funding to bladder function/health (28.1%) and SCI pathology/neuroprotection (25.1%). For the NIH, top categories included SCI regeneration/plasticity/repair (32.6%) and SCI pathology/neuroprotection (14.5%).

For awards employing exercise and rehabilitation-based interventions, the majority of funds were allocated to ambulation/locomotion (46.1%), SCI pain (13.8%), and upper limb/forelimb function (12.0%; **Fig. 12d**). The CDMRP and VA dedicated over half of their rehabilitation-related funding to ambulation/locomotion studies (57.2% and 53.4%, respectively), while the NIH allocated 28.3% to this category. Distinct agency-specific priorities also emerged: the CDMRP was the sole funder of rehabilitation-based psychological/psychosocial studies; the NIH was the only agency to support metabolism-focused studies in this category; and the VA exclusively funded studies targeting breathing, respiration, and cough. Together, these findings highlight the complementary and distinct priorities of federal agencies in addressing diverse aspects of SCI pathology through targeted therapeutic strategies.

## DISCUSSION

We curated and classified 1,589 federally funded SCI research awards from the NIH, CDMRP, and VA to systematically characterize how federal dollars have been allocated across the SCI research landscape. Using a structured schema that categorized awards across broad and diverse categories, we reveal striking differences in agency-specific funding priorities, thematic concentrations, and translational emphasis. These findings not only offer insights into how SCI research is currently structured and resourced in the United States but also serve as a foundation for future strategic planning, policy development, and community engagement.

### Complementary missions and funding patterns

Our analysis highlights the complementary roles that the NIH, CDMRP, and VA play within the broader SCI research ecosystem. The NIH, whose mission is “to seek fundamental knowledge about the nature and behavior of living systems and the application of that knowledge to enhance health, lengthen life, and reduce illness and disability”, predominantly supports basic and early-stage translational research. In contrast, the CDMRP portfolio is heavily oriented toward translational and clinical readiness, with particular emphasis on neuromodulatory device development and interventions targeting pain and bladder function, domains of critical relevance for those with SCI. This focus aligns with the CDMRP SCIRP’s stated goal of “[funding] research and [encouraging] multidisciplinary collaborations for the development and translation of more effective strategies to improve the health and well-being of Service members, Veterans, and other individuals with spinal cord injury”. Meanwhile, The VA research portfolio focuses on rehabilitation, muscle and bone health, and implementation studies that are directly aligned with the agency’s integrated healthcare delivery system. These priorities reflect the mission of the VA Office of Research and Development “to improve Veterans’ health and well-being via basic, translational, clinical, health services, and rehabilitative research”. While some overlap exists across agency portfolios, the differentiation in focus areas and translational stages appears to foster a more distributed and complementary approach to SCI research, potentially reducing redundancy and supporting a coordinated translational pipeline from discovery to implementation. It is also worth noting that the mechanisms for proposal solicitation vary among agencies. The NIH and VA typically announce general parent award mechanisms that do not dictate research themes; rather, the focus areas are determined by the applicants. In rare cases, research themes are predetermined by the NIH; e.g. the NIH HEAL Initiative, which has funded projects specifically centered on pain mechanisms and management (https://heal.nih.gov/). In contrast, the CDMRP SCIRP process invites proposals within specific areas of focus, which have historically included acute neuroprotection interventions, biomarker discovery, secondary health conditions, psychosocial issues, and rehabilitation/regeneration (https://cdmrp.health.mil/scirp/default). These divergent processes partially underscore the divergence in funding portfolios between the agencies.

### Justification for costs of SCI research spending

The World Health Organization (WHO) assigns a disability weight of 0.732 to cervical SCI, ranking it second only to acute schizophrenia (0.778) among 232 evaluated health states in terms of disability burden^7^. This high level of disability combined with a relatively normal life expectancy among individuals with SCI who survive past the acute phase results in a substantial long-term socioeconomic burden that far exceeds current federal investment in SCI research. According to our analyses, the federal government allocated $952 million to SCI research funding from 2017-2023, with a peak of $152 million in 2022 (**Fig. 3**). Phrased differently, those values represent investments by the government in developing treatments for SCI to relieve its burden on individuals and society. An in-depth analysis of the devastating long-term disability, decreases in quality of life, and mortality associated with SCI are beyond the scope of this analysis, and they cannot be measured in dollar values. However, the economic burden accrued by treatment costs and lost employment can be assessed, and in the US these costs far outstrip the investment made per year. The last published estimate of the total annual economic burden of SCI in the United States (dating back to 1997) was $7.74 billion^2^. Furthermore, a recent systematic review estimated that the lifetime cost per individual with SCI ranges from $0.7 million to $2.5 million^8^. Even using conservative assumptions, *the societal costs of SCI exceed federal research expenditures by at least 50-fold*, highlighting a critical discrepancy between the scale of the problem and the resources allocated to address it (See **Supplementary Discussion** for additional data). Previous research investigating the relationship between burden of disease and NIH fund allocations suggested that some diseases are historically overfunded, while others have received less attention^9^. The 2016 National Academies of Sciences, Engineering, and Medicine (NASEM) report on A National Trauma Care System: Integrating Military and Civilian Trauma Systems to Achieve Zero Preventable Deaths After Injury^10^ compiles reports and concerns about the systematic lack of research funds for traumatic injuries in general. Furthermore, the NASEM report also synthesizes previous reports. It provides recommendations such as: *“The committee recommends that funding for research on injury be commensurate with the importance of injury as the largest cause of death and disability of children and young adults in the United States”*.

Another important consideration in SCI funding decisions is the high cost of clinical trials. Depending on study design and participant requirements, the cost of conducting a single clinical trial can range from $3.4 million to $21.4 million, with a median per-participant cost estimated at $41,400^11,12^. In addition, the overall cost of developing a new therapeutic compound from discovery through clinical approval has been estimated to range from $1.31 billion to $2.3 billion^13,14^. These figures highlight the substantial financial investment required to advance SCI treatments from bench to bedside and suggest that the current scale of federal funding, while foundational, may be insufficient to fully support late-stage clinical translation and commercialization. Notably, private industry, particularly biopharmaceutical and medical device companies, plays a crucial role in bridging the translational gap between early-stage research (typically supported by NIH and CDMRP) and clinical implementation (supported by the VA). Industry is often responsible for the most expensive phase of the pipeline: late-stage development. For example, a single Phase 3 clinical trial funded by industry can cost upward of $100 million^15,16^, *a figure that approximates the entire annual NIH budget for SCI research*.

Growing recognition of the so-called “valley of death”, or the gap between promising preclinical data and commercially viable therapies, has spurred calls for targeted funding mechanisms to derisk innovation and attract private investment^17,18^. Additionally, nonprofit organizations, including patient advocacy groups and disease-specific foundations, also contribute across multiple phases of the research continuum, supporting pilot studies, infrastructure development, and translational research. It is a misconception that reductions in public funding for SCI care or research yield long-term cost savings. In reality, these expenses are often displaced to downstream systems such as Medicare and Medicaid, where the cost of long-term care for individuals with SCI remains substantial^1,2^. Given recent proposed changes in federal policy relating to Medicaid, the financial burden on individuals and their families could be increased. Increasing visibility into funding patterns, therapeutic pipelines, and patient needs is essential for maximizing return on investment and avoiding costly gaps or redundancies across the research and healthcare continuum.

### Recent and proposed changes in federal funding priorities

At the time of writing, substantial changes to the landscape of federal research funding and healthcare, both enacted and proposed, pose significant threats to the stability of the SCI research ecosystem. In March 2025, Congress chose to cease appropriations to the CDMRP SCIRP, resulting in an immediate loss of approximately $35 million annually in federal SCI research support. Proposed restructuring or elimination of the Administration for Community Living (ACL), which contains NIDILRR, threatens to lead to changes in funding and disruption of critical programs for individuals with disabilities. Additionally, in May 2025, the current administration proposed a federal budget that includes a ∼40% reduction in NIH funding (https://www.hhs.gov/about/budget/fy2026/index.html). While the specific impacts of this proposed NIH budget cut on SCI research are not yet known, a proportional decrease across all ICs would result in a 40% reduction in NIH SCI funding, equating to a loss of over $36 million based on FY2023 funding levels. Given the FY2023 SCI research expenditures ($91.4M from NIH, $35.7M from CDMRP, and $12.1M from VA), these combined changes could result in a total reduction of approximately 50% in federal SCI research support.

In consideration of the multi-year funding model used by the NIH, in which awards are paid out by installments on an annual basis, these proposed reductions in funding combined with the loss of SCIRP funding in FY25 will have an even larger impact on new research grants, affecting the already-falling NIH paylines. Such a dramatic contraction in funding would not only undermine ongoing research but could significantly destabilize the translational pipeline, particularly the clinical trial infrastructure, which has been heavily supported by CDMRP SCIRP funding. It is worth noting that these enacted and proposed funding cuts are occurring at a time of unprecedented progress in clinical SCI research; for example, the recently completed Up-Lift pivotal trial of a non-invasive spinal cord stimulation device for recovery of motor function (NCT04697472)showed positive safety and efficacy results which resulted in FDA de novo clearance (https://ir.onwd.com/static-files/0449dc19-a02d-4a81-aa0d-04051672e52d)^19^. More broadly, it has recently been predicted that proposed NIH budget cuts under the 47^th^ US presidential administration would severely hinder biomedical research, leading to an estimated $8.2 trillion in lost health value and $51 billion in economic losses, far outweighing the $500 billion in projected savings^20^.

The sudden loss of allocated funds reduces the amount available for SCI research, which, as described above, is already well below the required amount to move interventions through clinical trials and does not reflect the high burden of SCI. In light of these challenges, there is an urgent need to explore alternative sources of support, including enhanced investment from state agencies, private foundations, and public–private partnerships. Strategic collaborations such as TRACK-SCI^21,22^, which has fostered multicenter data harmonization and accelerated clinical insights, demonstrate how coordinated efforts can generate impactful outcomes even within constrained funding environments (https://cdmrp.health.mil/scirp/research_highlights/21track-sci_highlight). Our analysis provides a roadmap for identifying complementary strengths across agencies, recognizing funding gaps, and guiding more efficient and accountable investment of research dollars. Looking forward, sustaining progress in SCI research will require a more collaborative, transparent, and strategic funding infrastructure. It will also require clear communication among researchers, funders, policymakers, and people living with SCI to ensure that limited resources are allocated where they are most likely to advance meaningful outcomes.

### Alignment with research themes and community needs

Our classification framework provides insight into how federal SCI research funding is allocated across problem areas, interventions, and stages of readiness. Most federal awards continue to focus on motor and sensory function, regeneration, and neuroprotection, longstanding pillars of the SCI research landscape. While these areas represent vital scientific priorities with therapeutic potential, they do not always align with the lived experiences and quality-of-life concerns of individuals with SCI^23–26^. Encouragingly, bladder and urinary health, which are consistently ranked as top priorities by the SCI community^23,27^, have received substantial support, particularly from the NIH and CDMRP. However, other community-identified concerns remain vastly underfunded. Specifically, bowel and gastrointestinal health, autonomic dysfunction, sexual dysfunction, pressure sore injuries, and mental wellness have received relatively little federal investment despite their substantial impact on morbidity and mortality for those living with chronic SCI. Pain, another top concern, has received comparatively more attention, supported by both NIH and CDMRP, likely reflecting its broader relevance to other pathological conditions and significant socioeconomic burden. **Figure 13** provides a visual representation of how federal funds align with SCI community priorities, based on the responses from over 600 individuals conducted by the North American Spinal Cord Injury Consortium (NASCIC)^27^.

**Figure 13.**
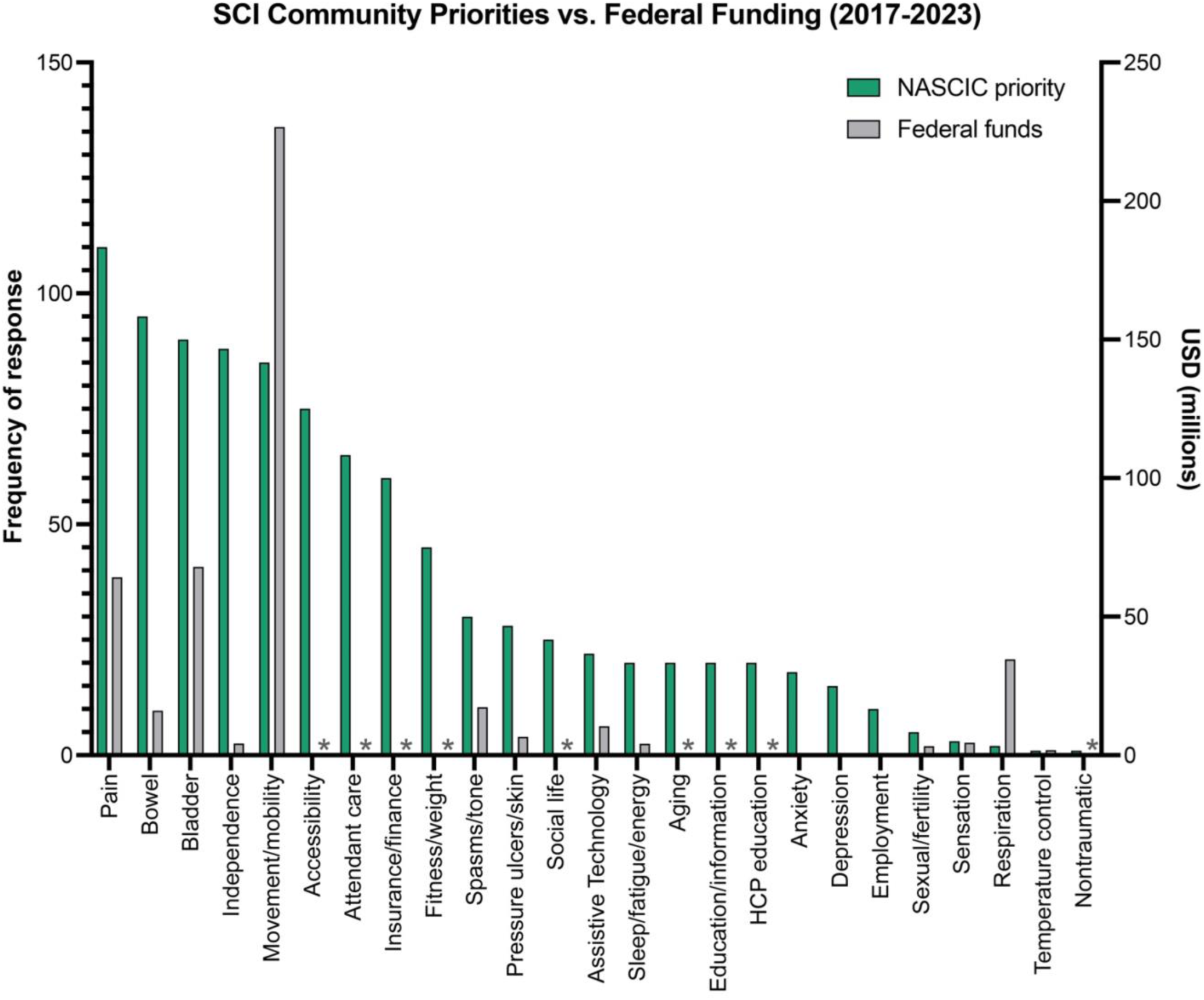
Comparison of SCI community priorities with federal research dollars. The green bars represent data from the 2019 North American Spinal Cord Injury Consortium (NASCIC) survey titled “Needs of the Community Living with SCI in North America”^27^, which asked a total of 644 adults living with SCI to answer the question, “What is the biggest challenge you face on a daily basis related to your spinal cord injury?”. Data indicates the number of responses in each category related to health, psychosocial, and socioeconomic issues. The gray bars represent the total amount of federal funding from NIH, CDMRP, and VA from the period of 2017- 2023 aligning with each category. Note that some issues captured in the NASCIC survey (e.g., accessibility), do not align with the categories used in our systematic analysis; these categories are indicated with asterisks.

The SCI2020 meeting highlighted the urgency of addressing these gaps^28^, yet our analysis suggests limited progress in shifting NIH funding toward secondary health conditions between 2020 and 2023. Despite decades of community advocacy and published surveys, the alignment between federal research priorities and the SCI community’s needs remains inconsistent. *One major contributing factor to this is that funding agencies can only fund the applications that they receive*. Investigators determine what research focus(es) to pursue, and there have been historically lower rates of applications in some high-priority areas, such as sexual function. There are promising models for change. For instance, NIH’s 2014 policy emphasizing sex as a biological variable significantly increased research funding and reporting on sex-based outcomes (https://orwh.od.nih.gov/sex-as-biological-variable). Other strategies, such as dedicated funding streams to incentivize work on underrepresented, high-impact secondary complications, such as pressure wounds and GI management over the lifetime, may help realign meaningful priorities. Allocating targeted funds for researchers branching into translationally relevant but understudied domains could accelerate progress in areas most meaningful to people living with SCI. Beyond working toward change at the level of federal funding agencies, these mechanisms could be implemented by private foundations and non-profit agencies, state governments, research institutions, and/or industry partners. This is a big problem to solve, and we propose the formation of working groups comprised of researchers, program staff, industry representatives, and individuals with lived SCI experience to focus on addressing and proposing solutions for adapting to critical losses in SCI research funding.

### Toward a living research map

A key outcome of this study is the proposal to develop a dynamic, publicly accessible platform to track and visualize federally funded SCI research across agencies. To date, there has been no unified view of SCI research funding classified by scientific objective, intervention type, or stage of translational readiness, nor by key indicators such as principal investigator, institution, or geography. A centralized, living dataset could fill this gap by enhancing transparency, standardizing classification systems, and enabling data-driven decision-making. The benefits of such a platform would be far-reaching. By offering a dashboard-style view of funding trends, gaps, and impacts, this could serve as a tool for researchers, funders, and community advocates alike. It would facilitate identifying redundant efforts and uncover synergistic opportunities, particularly among geographically proximate institutions working on complementary models (e.g., small animal, large animal, and human studies). Importantly, the platform could also support coordinated cross-agency funding strategies by visualizing shared priorities and investment trajectories over time.

This vision also aligns with recent collaborative initiatives in the SCI field that pool clinical and biological data across sites to enhance translational relevance. Similarly, clinical trial registries such as SCITrials.org and SCItrialfinder.net offer user-friendly portals for tracking therapeutic interventions—a model that could be emulated for research funding data. Looking ahead, such a platform could be integrated into existing community resources such as the ODC-SCI, or supported by professional societies such as the National Neurotrauma Society. We envision this platform as a shared infrastructure that supports interagency collaboration, fosters community engagement, and enables more strategic and accountable use of research dollars. Whether supported by public infrastructure (e.g., NIH, VA) or private foundations such as the CHNF, a research funding platform could serve as a catalyst to accelerate innovation and responsiveness in SCI research.

### Limitations

One limitation of our study is that it does not extend beyond the time period of 2008-2025 due to lack of publicly available data preceding 2008. Another significant limitation of our study is that it does not capture SCI research funding derived from agencies outside of the NIH, CDMRP, and VA. Therefore, we limit our work to US federal funding, which is an underestimate of the US funding ecosystem for SCI research. This omission is largely due to challenges in data availability and accessibility. For instance, while the CHNF is the largest private funder of SCI research in the United States—having awarded approximately $422 million from 2003 to 2024, or an average of $19 million annually—these data were not included in our analysis. This level of investment is comparable to the annual SCI research funding provided by the CDMRP (see **Figure 3**), positioning the CHNF as a major contributor to the SCI research landscape. Although grant data dating back to 2004 are publicly available on their Dimensions platform (https://chn.dimensions.ai/discover/grant), we elected not to include these data in the current study due to logistical constraints, including the inability to programmatically export and standardize thousands of awards. In the future, efforts to promote greater open communication and data sharing among federal and non-federal agencies could be utilized to promote standardized data reporting, similar to efforts by the Open Data Commons for SCI (ODC-SCI) to standardize research reporting ^29–31^.

In addition to the CHNF, other private funders such as the Wings for Life Foundation, the Paralyzed Veterans of America Research Foundation, the Praxis Institute, and the Craig Hospital Foundation support SCI research across the translational continuum, and these institutions prioritize patient-driven priorities. Other relevant government funding bodies, such as the National Institute on Disability, Independent Living, and Rehabilitation Research (NIDILRR; https://www.naric.com/pd-adv-srch), and state-level programs such as the New Jersey Commission on Spinal Cord Research and the New York State Department of Health Spinal Cord Injury Research Board, were also excluded due to lack of centralized or exportable funding data. Other DoD entities outside of CDMRP are not represented in this data set, including funding for SCI research efforts from the individual armed forces as well as DARPA; in addition, any SCI research funding from additional federal agencies are not included. While our analysis focuses on federally funded research with accessible and structured award data, future efforts that integrate private and state-level funding sources would provide a more comprehensive picture of the national SCI research ecosystem.

It is also likely that some NIH grants highly relevant to SCI were not captured in our current analysis. Specifically, there are awards that do not appear in the NIH RePORT “Spinal Cord Injury” dataset, despite their clear relevance to the field. For example, T32NS077889, titled “Neurobiology of CNS Injury and Repair”, is a training grant hosted at the University of Kentucky with a strong emphasis on SCI-related research and mentorship. However, it is not listed under the SCI portfolio on NIH RePORT, possibly due to limitations or inconsistencies in NIH’s internal tagging or portfolio classification processes. The criteria used by NIH to determine which awards are included in disease-specific portfolios are not publicly defined, making it difficult to systematically address such omissions. Complicating things further, the concept of “relevance” to SCI is inherently subjective. For instance, a study focused on transcriptional mechanisms of spinal cord development may be considered relevant by basic SCI researchers, while individuals with lived experience may view it as too distant from clinical application to be considered SCI research. In reality, SCI relevance exists along a continuum, from foundational mechanistic studies in intact systems to direct interventions in injured populations. For example, advancing our understanding of bladder dysfunction after SCI will likely depend on better understanding the neural control of the bladder in the uninjured nervous system. To avoid introducing subjective bias into our classification process, we elected to rely on NIH’s own designation of SCI-relevant awards as reported in the RePORT “Spinal Cord Injury” portfolio. While this approach may exclude some grants that investigators or community members would consider relevant, it provides a transparent and reproducible foundation for analysis.

## CONCLUSION

Federal funding for SCI research is broadly aligned with complementary agency missions, but critical gaps remain in matching community priorities, supporting infrastructure, and advancing translational research. There is a pressing need to reduce redundancy in SCI research while increasing representation in critically underfunded areas that align with community priorities. A centralized, living research dashboard would serve as a vital tool for the SCI ecosystem—promoting efficiency, collaboration, and ultimately, progress toward improving quality of life for individuals living with SCI. We propose the development of a working group to focus on addressing and proposing solutions for adapting to critical losses in SCI research funding.

## Data Availability

All data produced in the present work are contained in the manuscript

## ABBREVIATIONS

CDMRP: Congressionally Directed Medical Research Program
CHNF: Craig H. Neilsen Foundation
DoD: Department of Defense
DTIC: Defense Technical Information Center
FY: fiscal year
IC: Institute or Center
NASCIC: North American Spinal Cord Injury Consortium
NIH: National Institutes of Health
ODC-SCI: Open Data Commons for Spinal Cord Injury
RePORT: Research Portfolio Online Reporting Tools
RePORTER: Research Portfolio Online Reporting Tools Expenditures and Results
SCI: spinal cord injury
SCIRP: CDMRP Spinal Cord Injury Research Program
TRACK-SCI: Transforming Research and Clinical Knowledge in Spinal Cord Injury
WHO: World Health Organization
VA: Department of Veterans Affairs

## ACKNOWLEDGMENTS

We gratefully acknowledge Lyn Jakeman (NINDS), Carol Taylor-Burds (NINDS), Linda Bambrick (SCIRP), Timothy Brindle (VA-RRD), and Naomi Kleitman (CHNF), who developed the three-axis classification system that we modified and used in our study. We thank Marco Sorani, Melissa Miller (SCIRP), Adam Ferguson, and Prakruthi Amar Kumar for helpful discussion, and John Houle for providing abstracts for P02NS055976. This work was supported in part by the National Institutes of Health (R01NS116404 to J. N. Dulin, R35GM138098 to H. Blackmon, R01NS122961 to D. A. McCreedy), the Craig H. Neilsen Foundation (#546639 to J. N. Dulin), Mission Connect, a program of TIRR Foundation (#021-101 to J. N. Dulin), the Paralyzed Veterans of America Research Foundation (#3195 to B. R. Kondiles, #3196 to R. N. Malik), the National Science Foundation Graduate Research Fellowships Program (DGE2139772 to M. R. Pacheco, DGE2140743 to J. Biundo), Michael Smith Health Research BC (CTTP-2024-04574 to R. N. Malik), and the Canadian Training Platform for Trials Leveraging Existing Networks (CANTAP-2023-03855 to R. N. Malik).

