## Supplementary Information for "Funding distributions, trends, gaps, and policy implications for spinal cord injury research: A systematic analysis of US federal funds"

---

### SUPPLEMENTARY METHODS

*Three-axis classification schema*

The System/Problem axis contains general categories that describe where an award falls within the overall scope of SCI research questions, as well as subcategories within each of the general categories. "SCI Adjacent" comprises any awards that do not directly focus on SCI but instead are judged to be indirectly relevant to SCI research. Examples include studies of the intact spinal cord (e.g., NIH ZIANS003153, which seeks to understand how intact spinal cord circuits encode neural programs for motor behavior); studies focused on characterizing the physiology of SCI-relevant body systems (e.g., NIH R01HL080209, which seeks to characterize mechanisms of respiratory plasticity in the intact body); and studies to develop novel tools and/or technology relevant to SCI (e.g., NIH R01NS090617, which aims to develop tools for isolation and culture of ventral spinal neuron populations). "SCI Pathology & Repair" encompasses awards that are either focused on SCI pathology and/or neuroprotective mechanisms, or SCI regeneration/repair and plasticity, but do not primarily evaluate functional outcomes. Examples include NIH R01NS122961, which focuses on characterizing the effect of diclofenac treatment on neutrophil activation and tissue sparing; and NIH R01NS123708, which aims to define the cellular and molecular mechanisms of glial bridging in spinal cord regeneration. Studies that are primarily focused on evaluating functional and/or physiological outcomes following SCI fall into the "SCI Motor/Sensory Function" and "SCI Secondary Health Conditions" categories. There are 22 System/Problem subcategories encompassing specific functional/physiological outcomes such as upper limb/forelimb function (e.g., CDMRP W81XWH-21-1-0551, which aims to improve arm and hand function for individuals with chronic cervical SCI through transcutaneous stimulation); bladder/urinary health (e.g., CDMRP HT9425-23-1-0522, which utilizes chemodenervation of the bladder to combat neurogenic bladder after SCI in rodents); SCI pain (e.g., CDMRP W81XWH-22-1-0522, which seeks to evaluate the contributions of opioids to long-term pain symptoms in rodent SCI models); and muscle & bone health (e.g., VA I01RX002089, which employs low-intensity whole body vibration to prevent bone loss after SCI in rodents). Additional categories within the System/Problem axis include "Acute SCI Management", encompassing studies focused on emergency management of SCI; "Sociological", encompassing studies focused on psychosocial, sociological, or community living aspects of SCI; and "Infrastructure", which captures research and/or clinical resource awards to support SCI research, clinical centers and network hubs, and support for training, conferences, and equipment. "Other SCI studies" is an orphan category that includes studies relevant to SCI that do not fall into any of the above categories.

The Approach/Intervention axis contains categories describing whether and what type of therapeutic intervention(s) are used in the study. Studies not utilizing therapeutic interventions can be classified as either "Discovery"-based or "Tools and Technology"-based. Such awards include *in vitro* or *ex vivo* modeling studies (e.g., NIH F30NS061403, which studies mechanisms of axon growth inhibition *in vitro*); *in vivo* studies that do not utilize interventions (e.g., NIH R01DK126008, which seeks to characterize the mechanisms underlying liver pathology in SCI); human clinical observational studies (e.g., CDMRP W81XWH-16-1-0497, which seeks to identify the critical care variables that predict sensorimotor and autonomic outcomes after SCI in human subjects); studies focused on development of diagnostic tools or biomarker discovery (e.g., VA I01RX003081, which aims to identify biomarkers of pressure injury risk in persons with SCI); and studies that focus on development of sensors, activity monitors, or computer interfaces (e.g., CDMRP W81XWH-16-1-0602, which seeks to develop near-infrared spectroscopy as a sensor to monitor blood oxygenation and perfusion following SCI). Studies utilizing therapeutic interventions fall into one of 11 subcategories according to the type of intervention used. Examples include surgical interventions (e.g., CDMRP W81XWH-15-1-0456, a clinical trial to evaluate efficacy of nerve transfers for treatment of cervical SCI in humans); drug interventions (e.g., VA I01RX003986, which aims to test an iron chelator drug for safety and efficacy in rodent SCI); cell therapies (e.g., NIH R01NS116404, which seeks to evaluate the efficacy of neural progenitor cell transplantation in rodent SCI); neuromodulatory devices (e.g., CDMRP HT9425-24-1-0218, which aims to evaluate efficacy of electrical stimulation to promote forelimb functional recovery in rats and humans); lifestyle-based interventions (e.g., VA I01RX002116, which tests the efficacy of a program to improve adherence to positive airway pressure therapy in SCI subjects with sleep-disordered breathing); and others. Finally, additional awards in the "Resources/Other" category may support community programs, conferences, research and/or clinical resources, training, and data management.

The Readiness axis describes the scope of the award on the translational axis; i.e., where does the study fall on the spectrum of basic research to advanced clinical studies. In this axis, "Basic-basic" research constitutes basic research studies that do not include injury models or therapeutic interventions. As an example, NIH grant R01NS044916 explores the molecular mechanisms underlying formation of structures associated with the nodes of Ranvier. "Disease-related basic" research awards include studies that utilize SCI models but no therapeutic intervention (e.g., NIH R01NS041548, which aims to characterize how noxious stimulation underlies plasticity in a rodent spinal contusion model). "Preclinical" research encompasses animal studies that utilize therapeutic interventions (e.g., VA I01RX002483, which assesses the effects of plasticity-promoting strategies on corticospinal regeneration in mice after SCI). Clinical studies were subcategorized into Phase 0 or early feasibility studies (e.g., CDMRP HT9425-23-1-0522, which includes a pilot clinical trial to examine the effects of botulinum toxin A on bladder compliance); Phase I or II trials (e.g., CDMRP W81XWH-14-2-0193, which comprises a 2-year double-blinded, randomized, controlled trial, comparing the effects of zoledronic acid to placebo on bone health); and Phase III or pivotal trials. Only one Phase III clinical trial was identified in our dataset: CDMRP W81XWH-16-1-0748, a Phase III multicenter, randomized, controlled study to investigate the efficacy and safety of a blood pressure augmentation strategy in patients with acute SCI. A complete list of all categories in each of the three axes is provided in **Table 1**.

*Statistical analysis of combinatorial approaches*

To explore patterns of combinatorial therapeutic approaches in federally funded SCI research (**Figure 8c; Table 3**), we analyzed how often different intervention types were co-utilized within individual awards. For this analysis, projects classified as combinatorial (i.e., involving more than one therapeutic approach) were examined. Intervention pairs were represented as an undirected network, with each node corresponding to a distinct therapeutic category and each edge representing the frequency with which that pair of interventions appeared together across projects. To evaluate whether specific combinations occurred more frequently than expected by chance, we calculated the expected frequency of each pair under an independence model based on the marginal frequencies of each intervention type. Statistical significance was assessed using one-tailed Poisson tests, with Bonferroni correction applied for multiple comparisons (47 tests; the number of unique pairings in the dataset). Network visualization was performed in R Studio using a custom script. In the resulting network diagram (**Fig. 8c**), edges are scaled by the observed frequency of co-occurrence, with solid lines reflecting statistical significance ($P < 0.05$, corrected), nonsignificant edges are shown as dashed lines.

### SUPPLEMENTARY DISCUSSION

*Cost of SCI to the individual and society*

Compiled data updated for 2023 dollar equivalents by the Spinal Cord Injury Model Systems Knowledge Translation Center estimates expenses ranging from ~$429,000 - $1,315,000 per person in the first year after SCI[1]. Costs for each subsequent year range from ~$52,000 - $228,000 per year. These estimates fail to include indirect costs such as lost wages, which are estimated at $92,578 dollars per year[1]. Roughly half of people suffering an SCI have private insurance. Assuming the percentages of healthcare costs paid by private insurance, government insurance, or out of pocket approximately match those seen across all healthcare spending in the US[2], government health insurance covers 42.6% of the cost[3].

While these estimates and assumptions are not exact, they are helpful for contextualizing the healthcare costs paid by the government for SCI compared to its investment in SCI research. If 1,000 individuals sustain an SCI requiring the lowest estimated healthcare costs for the first year post-injury of $440,000, costs paid by governmental insurance equal almost $190 million (42.6% of $440 million). This conservative estimate greatly exceeds the amount allocated by the government at the peak funding year described in this study. If only 500 people sustaining injuries requiring the highest estimate of costs in the first year, they would still more than double the peak funding provided by the government (42.6% of the $1.37 million total cost in the first year post-injury for 500 individuals = $291 million.) Using the latest incidence for SCI in the US of 18,000 cases, and assuming each requires the average first-year costs, government insurance pays $5.33 billion per year. These estimates fail to consider any costs paid by governmental insurance for the remainder of the person's lifetime. SCI research has the potential to reduce healthcare cost by improving treatments, reducing hospital stays, preventing/treating secondary complications, and reducing rehospitalizations. Even the most conservative estimated values demonstrate the power of investing in SCI research for reducing later economic burden for individuals and the government.

Of course, these hypotheticals neglect the complexities of prioritizing federal funding allocations to reflect multi-faceted and ever-changing priorities. Furthermore, different agencies are responsible for research and government insurance. Subsequently, it is also appropriate to consider our findings in the context of the funding landscape and burdens of different pathologies. Our analysis found that NIH spent $91.4 million on SCI research (**Fig. 3**), 0.15% of NIH's reported 2023 funding of $49.18 billion (https://www.nih.gov/about-nih/organization/budget). In 2023 NINDS spent $2.58 billion on research (including extramural and intramural, excluding SBIR/STTR small business grants; https://www.ninds.nih.gov/about-ninds/what-we-do/budget-legislation/fy23-total-appropriation). Our analysis shows that in 2023, NINDS allocated $74.8 million to SCI research, comprising less than 3% of its research expenditure. We highlight NINDS, as the disease burden of SCI is frequently contextualized within neurological disorders[4]. Of the 14 disorders listed, SCI had the 7$^{th}$ highest incidence and 5$^{th}$ highest prevalence in the US in 2017; unfortunately, mortality rates and disability adjusted life years were not available. Doubtless, advancing research for all neurological diseases is important, and it is not our intention to set priorities for research allocations. It is worth noting that in 2023, 16 of the NINDS grants analyzed, amounting to $9,200,000, were classified as infrastructure or SCI-adjacent, implying benefits from these grants generalize beyond SCI. A rising tide raises all boats, and increasing funding for neurological disorder research alongside increasing transparency on priorities and current usage will enable researchers and funding agencies to provide the highest return on investment for research dollars.

### SUPPLEMENTARY TABLE LEGENDS

**Supplementary Table 1.** All awards in the NIH RePORT "Spinal Cord Injury" funding portfolio from 2008-2023.

**Supplementary Table 2.** NIH Institutes and Centers.

**Supplementary Table 3.** NIH spinal cord injury research awards from the 2008-2023 RePORT "Spinal Cord Injury" portfolio annotated with our classifications in columns AK-AU.

**Supplementary Table 4.** NIH research awards from the 2008-2023 RePORT "Spinal Cord Injury" portfolio that were excluded from our analysis.

**Supplementary Table 5.** CDMRP SCIRP spinal cord injury research awards from 2009-2023 annotated with our classifications in columns L-O.

**Supplementary Table 6.** VA spinal cord injury research awards from 2017-2025 annotated with our classifications in columns AC-AG.

### SUPPLEMENTARY FIGURES

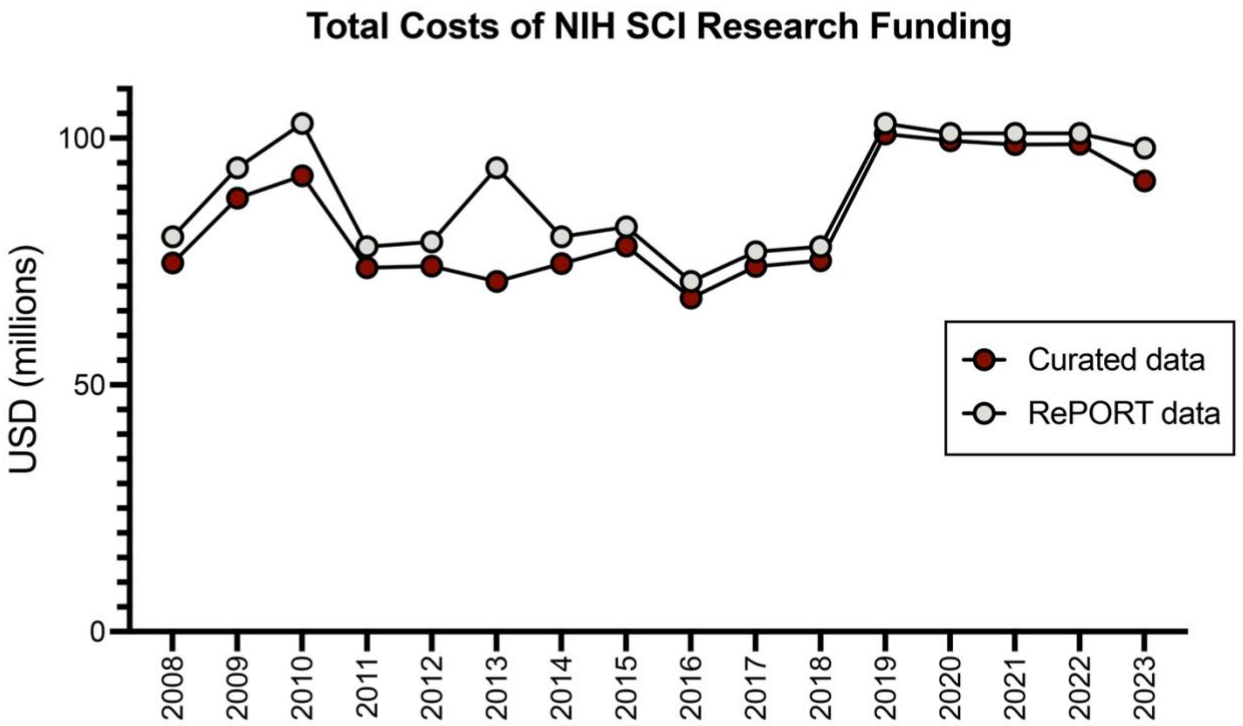


**Supplementary Figure 1.** Comparison of total annual costs of NIH spinal cord injury research awards from RePORT "Spinal cord injury" portfolio (gray) versus our curated dataset excluding irrelevant awards (maroon).

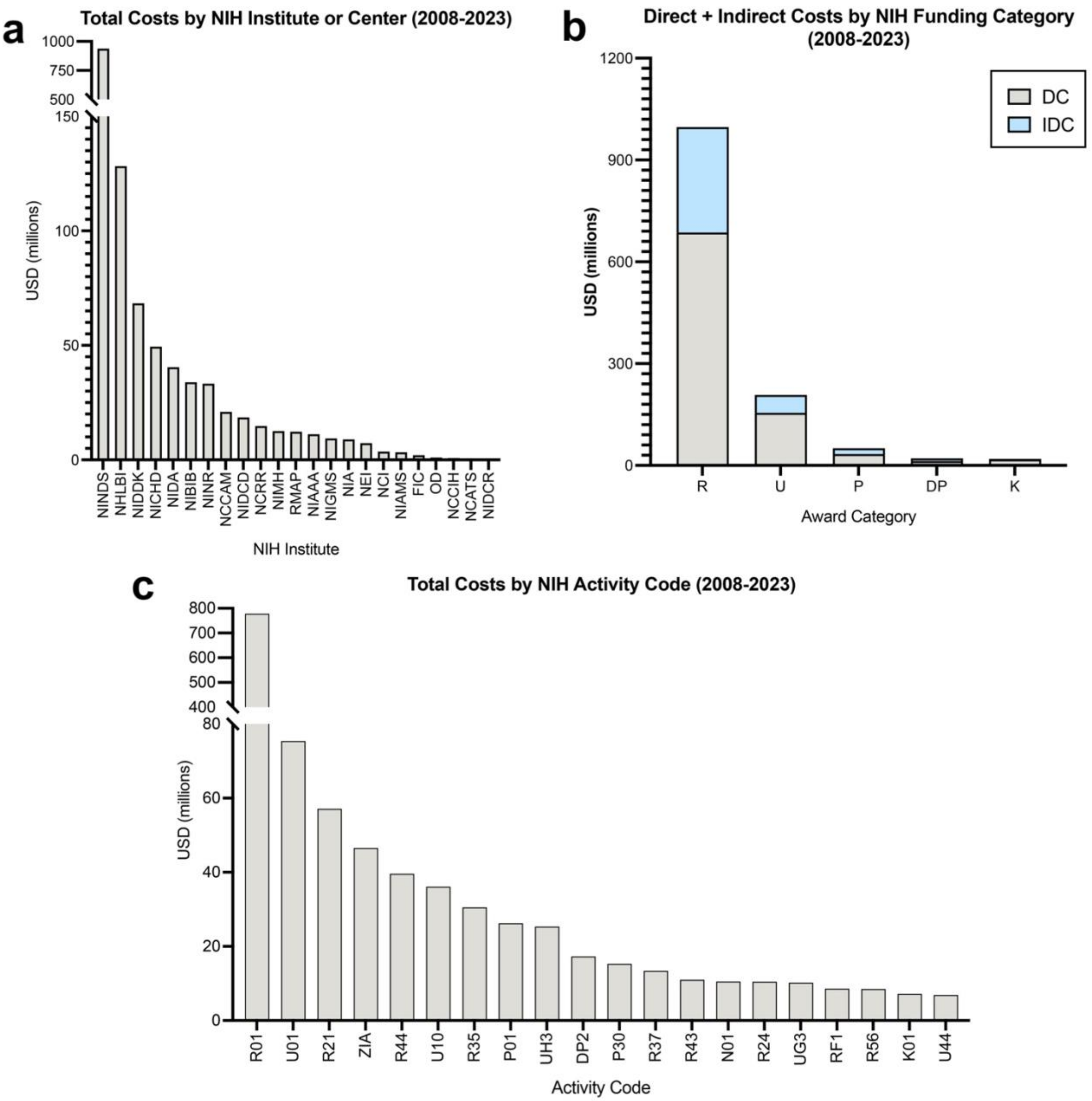


**Supplementary Figure 2. Breakdown of NIH spinal cord injury research awards, 2008-2023.** (**a**) Funds allocated to individual NIH Institutes and Centers. FIC, Fogarty International Center; NCATS, National Center for Advancing Translational Sciences; NCCIH/NCCAM, National Center for Complementary and Integrative Health; NCI, National Cancer Institute; NCRR, National Center for Research Resources*; NEI, National Eye Institute; NHLBI: National Heart, Lung, and Blood Institute; NIA, National Institute on Aging; NIAAA, National Institute on Alcohol Abuse and Alcoholism; NIAMS, National Institute of Arthritis and Musculoskeletal and Skin Diseases; NIBIB, National Institute of Biomedical Imaging and Bioengineering; NICHD, Eunice Kennedy Shriver National Institute of Child Health and Human Development; NIDA, National Institute on Drug Abuse; NIDCD, National Institute on Deafness and Other Communication Disorders; NIDCR, National Institute of Dental and Craniofacial Research; NIDDK, National Institute of Diabetes and Digestive and Kidney Diseases; NIGMS, National Institute of General Medical Sciences; NIMH; National Institute of Mental Health; NINDS, National Institute of Neurological Disorders and Stroke; NINR, National Institute of Nursing Research; OD, NIH Office of the Director; RMAP, NIH Roadmap for Medical Research*. *Indicates that NIH Institute or Center is no longer active. (**b**) Direct costs (DC) and indirect costs (IDC) stratified by top 5 NIH research funding categories. DP, Director's Pioneer awards; K, individual career development; P, program grants; R, research grants; U, cooperative agreements. (**c**) Total costs of NIH awards within the top 20 activity codes.

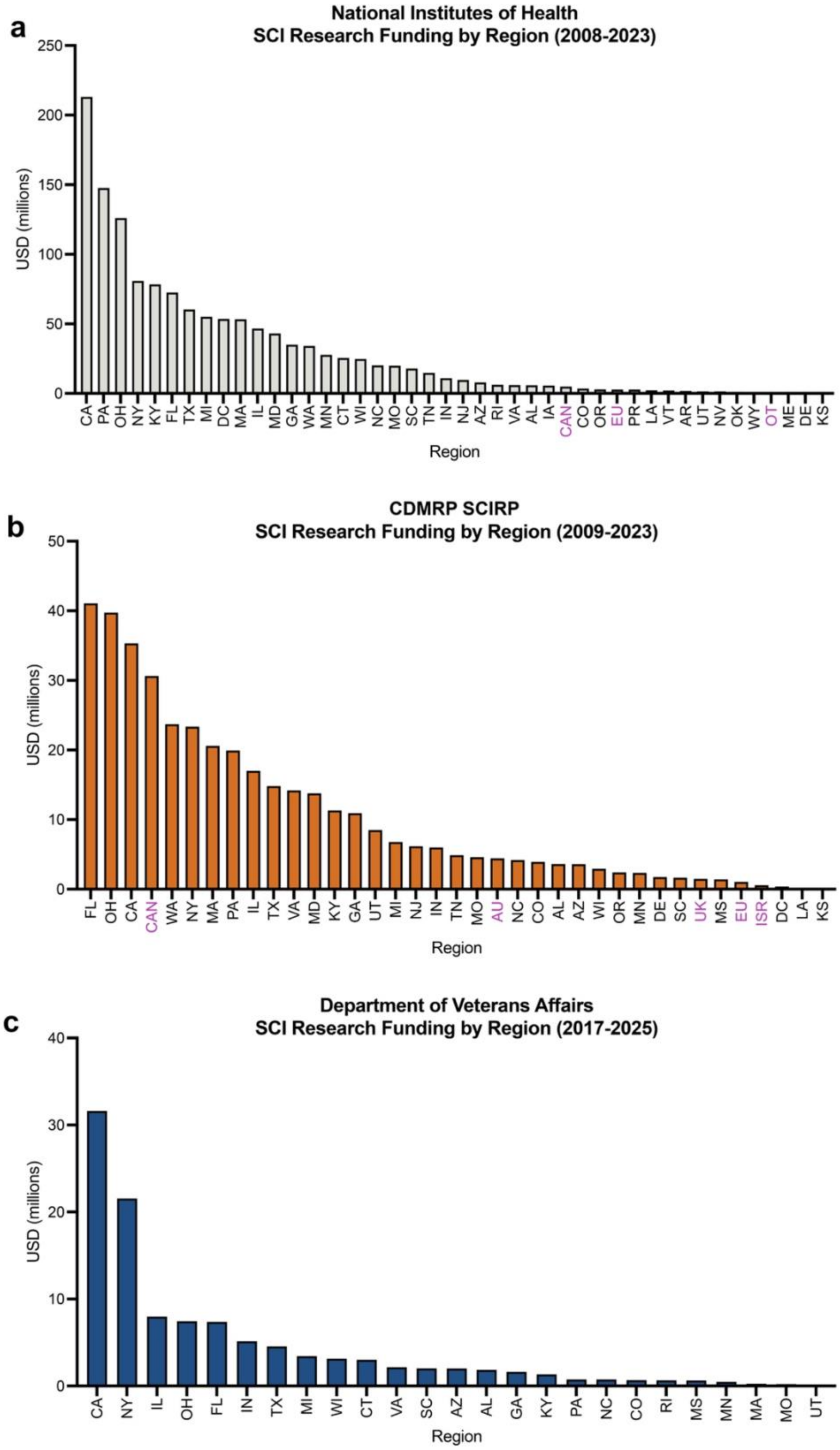


**Supplementary Figure 3.** (**a-c**) Federal funds allocated to each U.S. state or regional entity by (**a**) NIH, (**b**) CDMRP, and (**c**) VA. Non-U.S. entities are labeled in magenta: AU, Australia; CAN, Canada; EU, European Union; ISR, Israel; OT, other; UK, United Kingdom. For each graph, states that did not receive any federal funding are not shown.

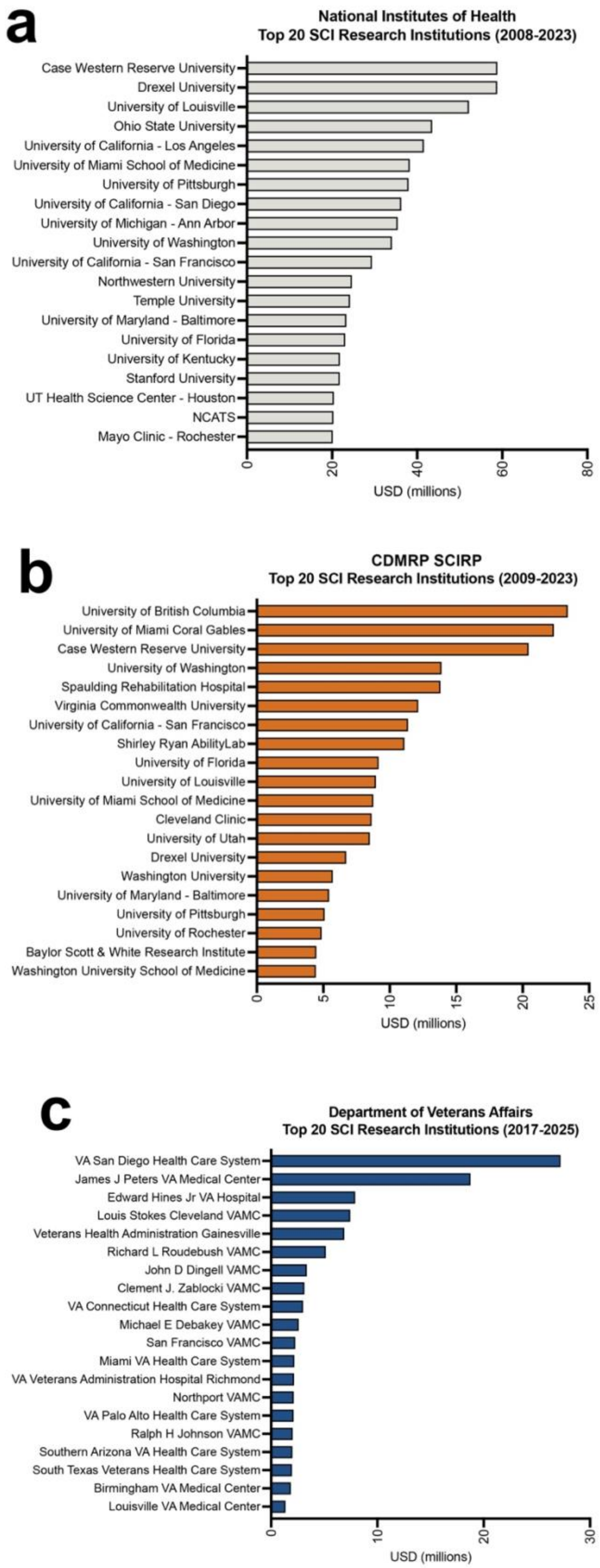


**Supplementary Figure 4. Top awarded institutions.** (**a**) Top 20 institutions receiving SCI research funding from the NIH, 2008-2023. (**b**) Top 20 institutions receiving SCI research funding from the CDMRP, 2009-2023. (**c**) Top 20 institutions receiving SCI research funding from the VA, 2017-2025.

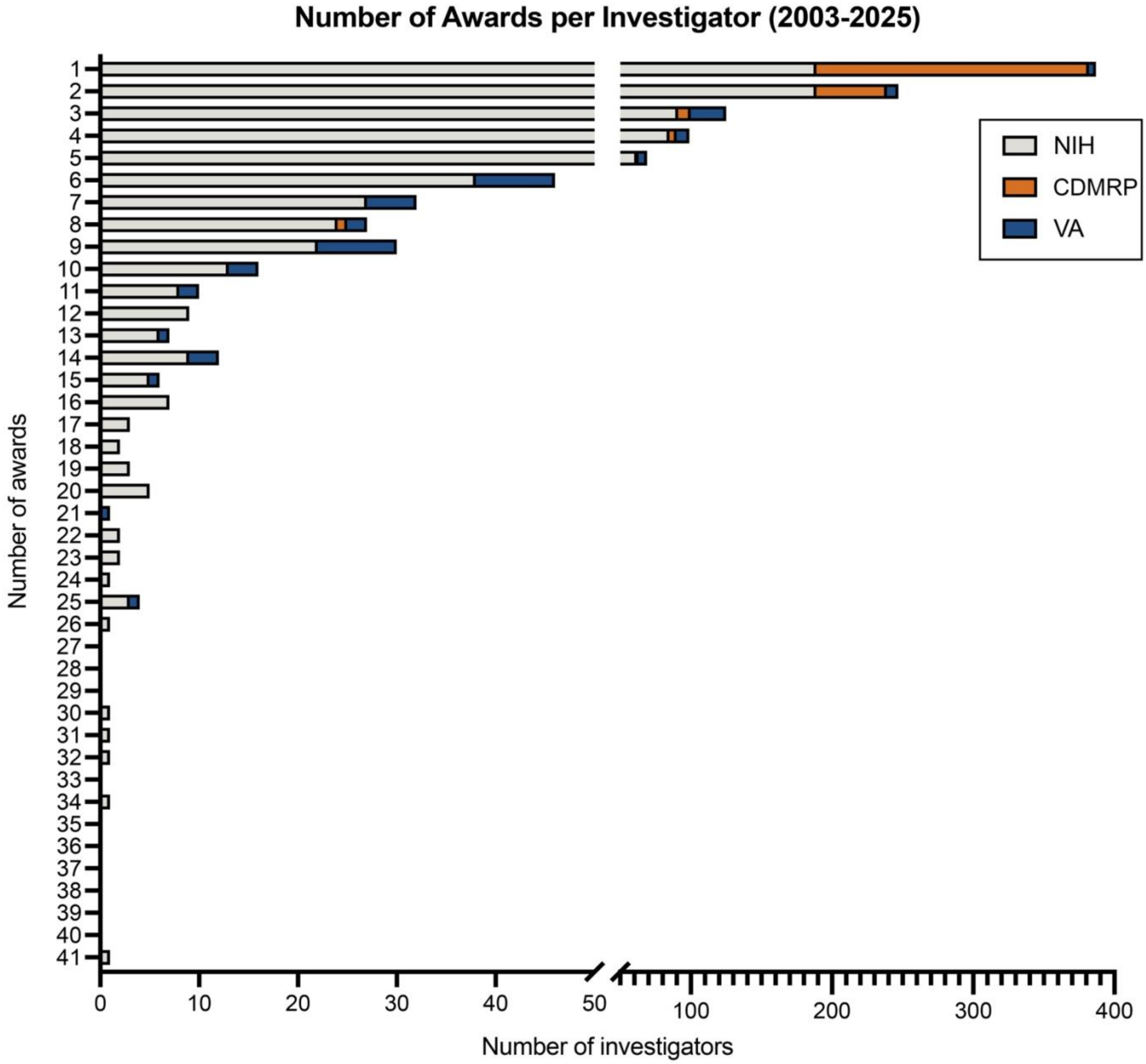


**Supplemental Figure 5. Frequency of awards to individual investigators.** Data for the NIH (2008-2023), CDMRP (2009-2023), and VA (2017-2025) are presented.

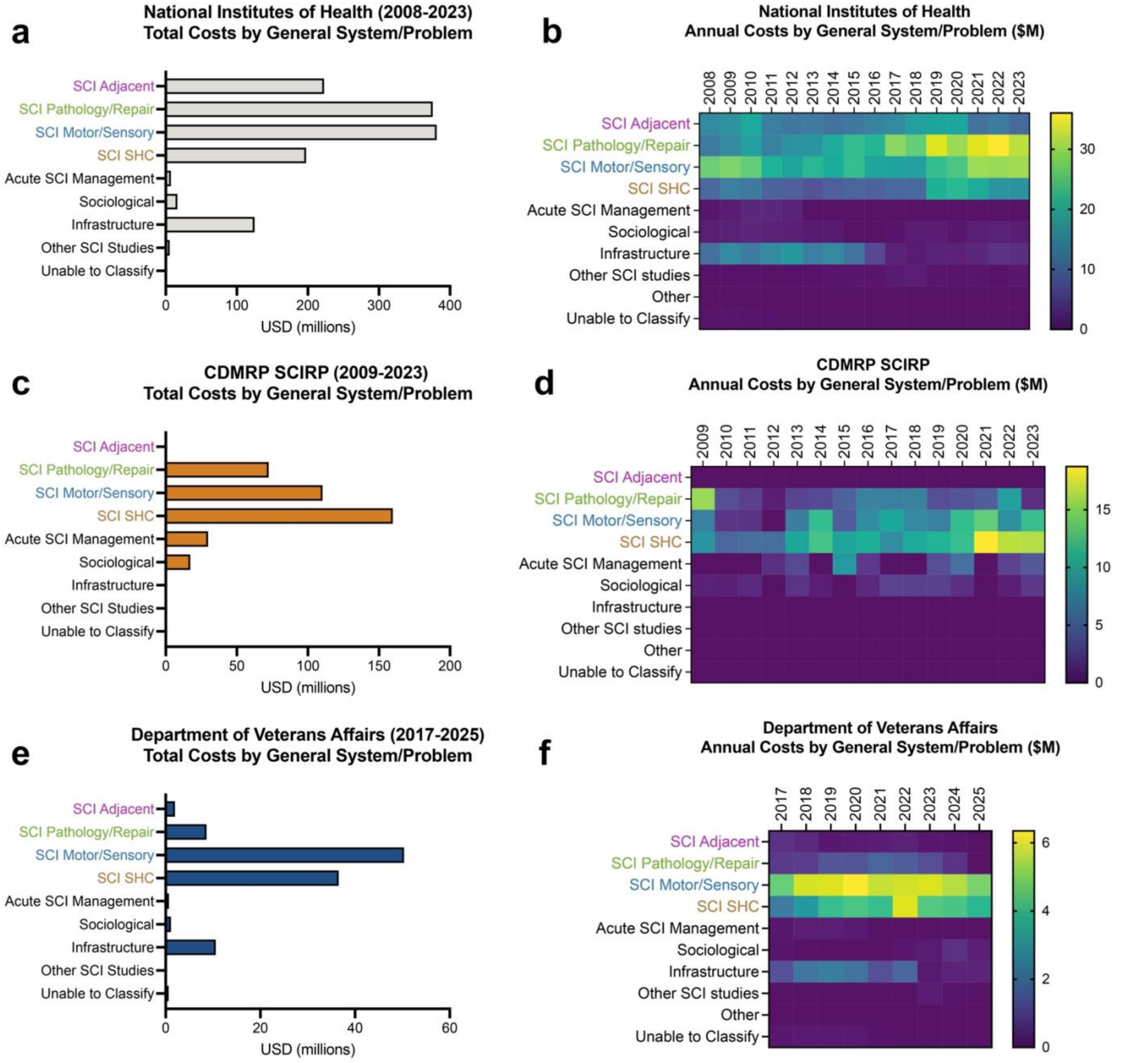


**Supplementary Figure 6. Federal SCI research awards classified by general system/problem category.** Total costs of SCI research awards classified by general system/problem category for (**a, b**) NIH from 2008-2023, (**c, d**) CDMRP from 2009-2023, and (**e, f**) VA from 2017-2025. (**a, c, e**) Total costs in millions of dollars for the entire analyzed time period. (**b, d, f**) Annual costs; color scales are in millions of dollars.

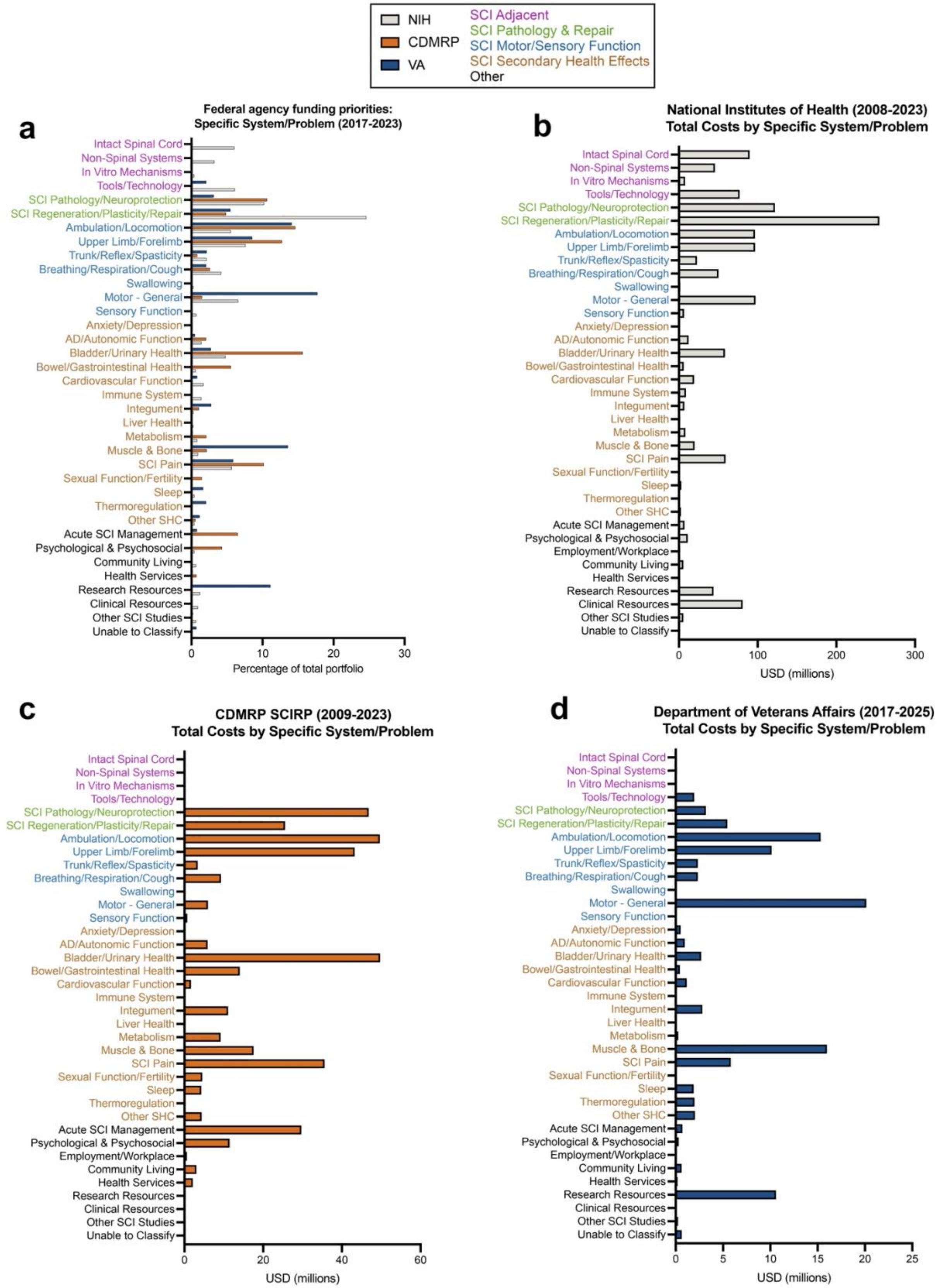


**Supplementary Figure 7. Total federal spending by specific system/problem category.** (**a**) Proportions of each federal agency's budget allocated to each category; data are expressed as a proportion of 100% for each agency. (**b-d**) Total costs of SCI research awards classified by specific system/problem category for (**a**) NIH from 2008-2023, (**b**) CDMRP from 2009-2023, and (**c**) VA from 2017-2025. Specific categories are color-coded according to the general system/problem category.

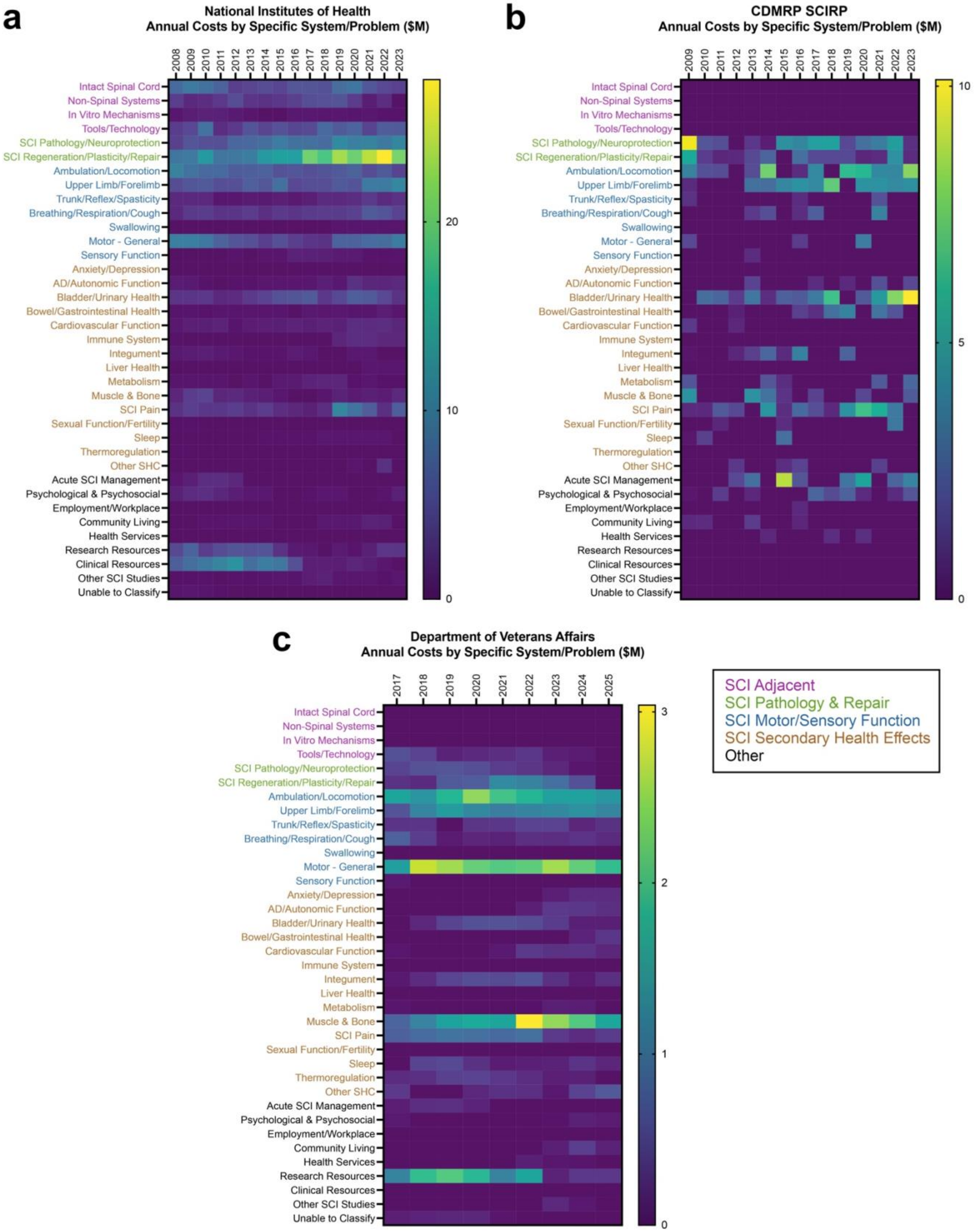


**Supplementary Figure 8. Total federal spending over time by specific system/problem category.** Total annual costs of SCI research awards classified by specific system/problem category for (**a**) NIH from 2008-2023, (**b**) CDMRP from 2009-2023, and (**c**) VA from 2017-2025. Specific categories are color-coded according to the general system/problem category.

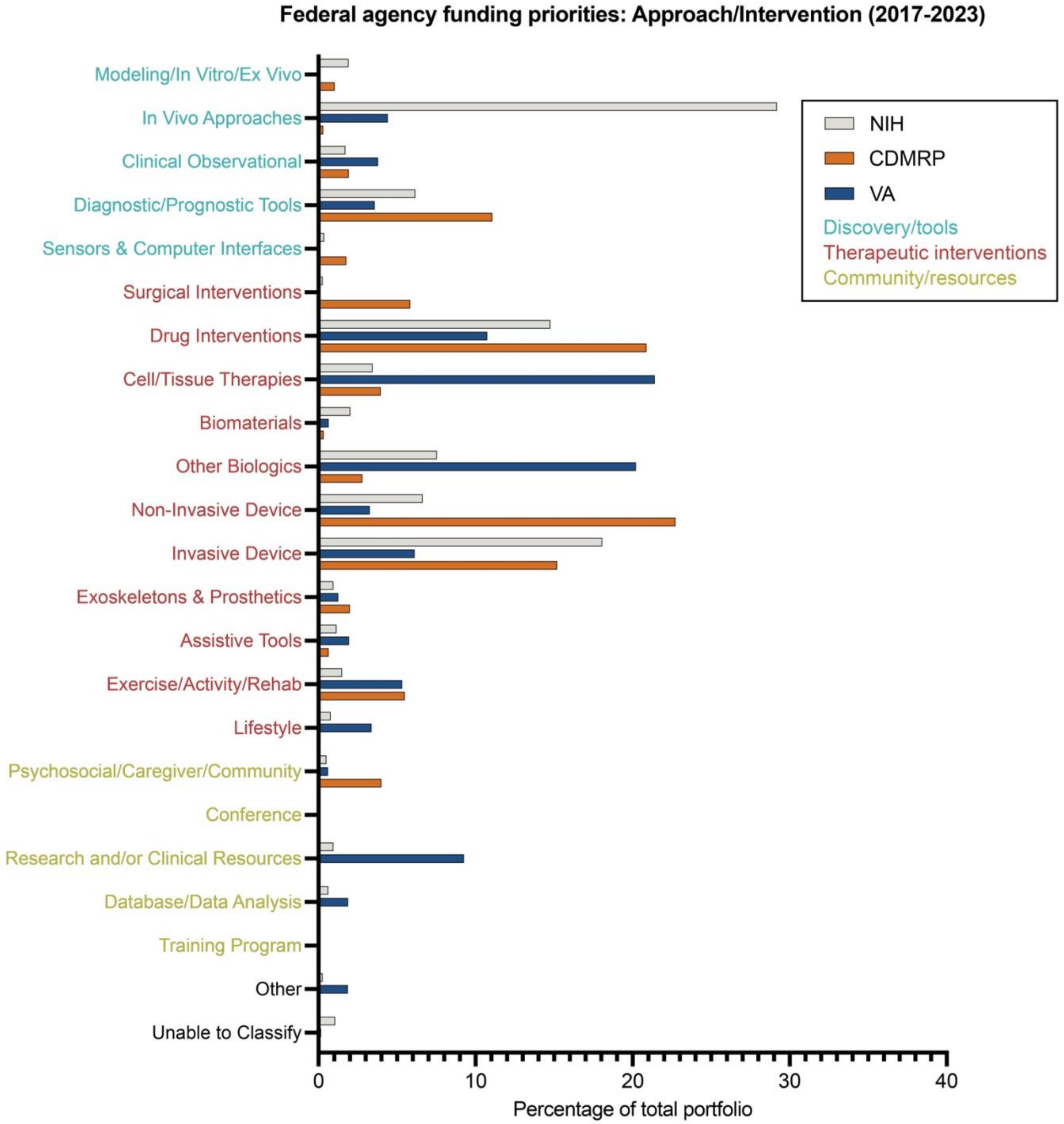


**Supplementary Figure 9.** Proportions of each federal agency's budget allocated to each approach/intervention category; data are expressed as a proportion of 100% for each agency.

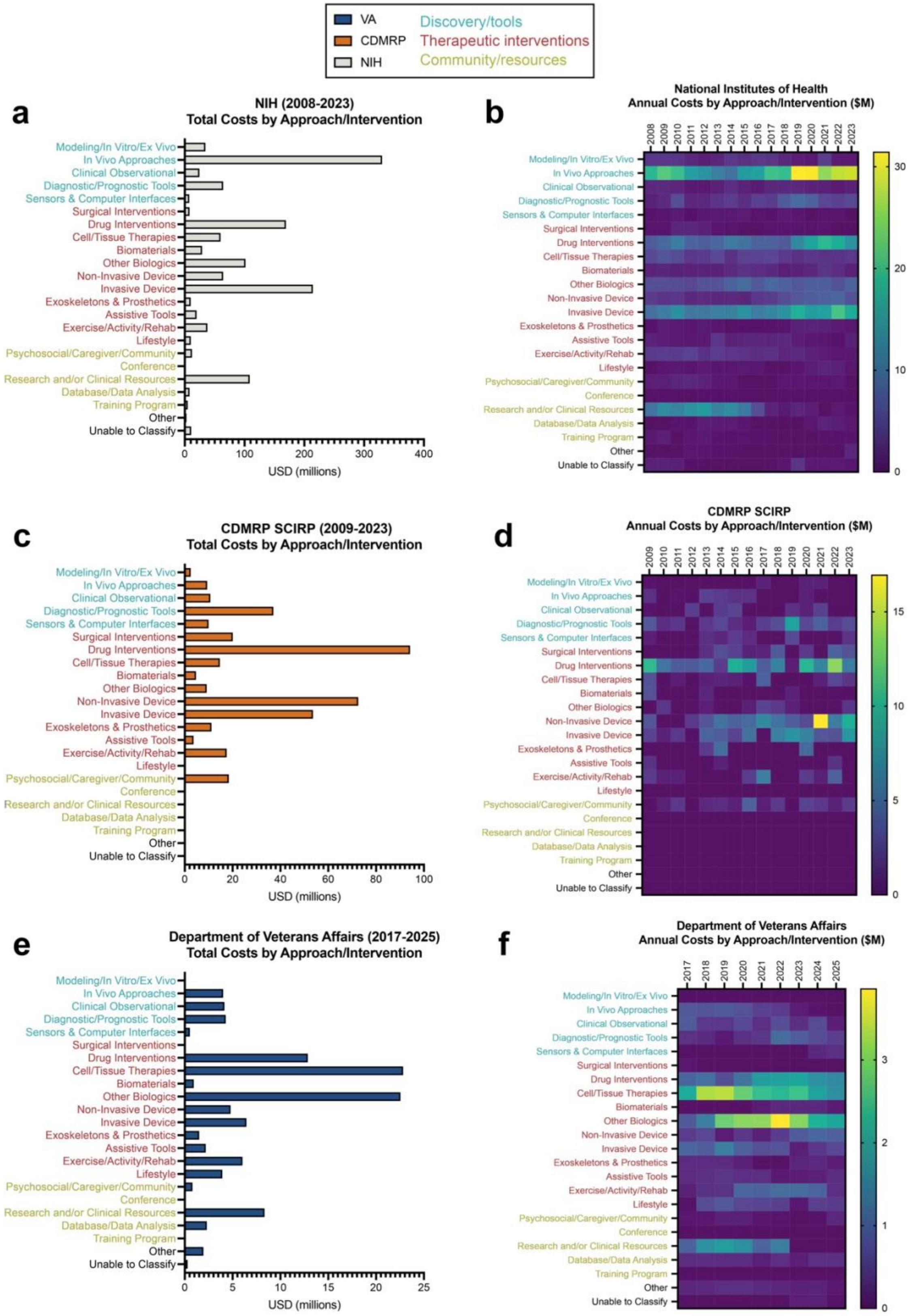


**Supplementary Figure 10. Total federal spending by approach/intervention category.** Total costs of SCI research awards classified by approach/intervention category for (**a, b**) NIH from 2008-2023, (**c, d**) CDMRP from 2009-2023, and (**e, f**) VA from 2017-2025. (**a, c, e**) Total costs in millions of dollars for the entire analyzed time period. (**b, d, f**) Annual costs; color scales are in millions of dollars. Specific categories are color-coded according to the class of approach or intervention used.

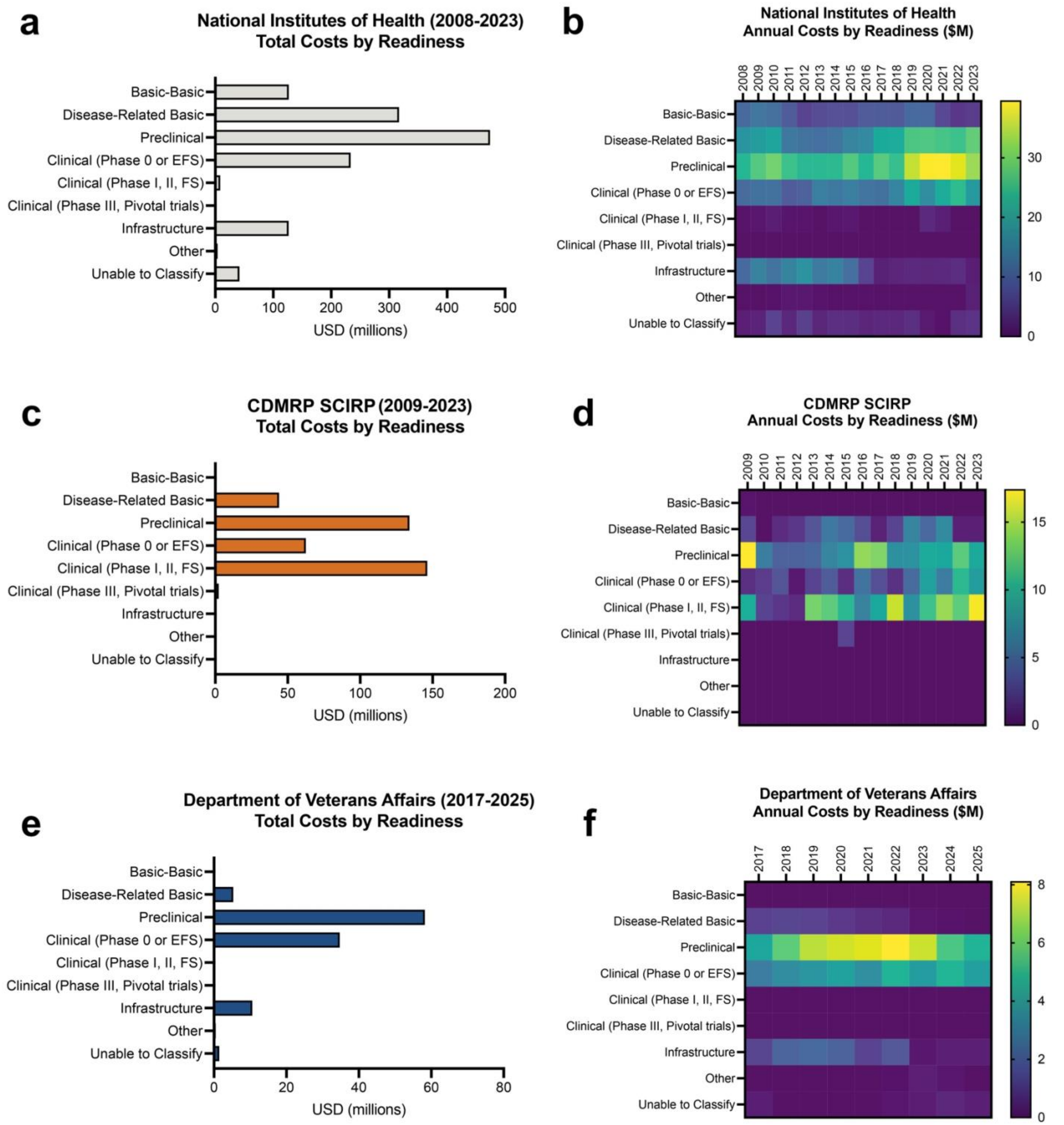


**Supplementary Figure 11. Total federal spending by readiness category.** Total costs of SCI research awards classified by readiness category for (**a, b**) NIH from 2008-2023, (**c, d**) CDMRP from 2009-2023, and (**e, f**) VA from 2017-2025. (**a, c, e**) Total costs in millions of dollars for the entire analyzed time period. (**b, d, f**) Annual costs; color scales are in millions of dollars.
